# Transcriptomic and Genomic Sequencing of South African Colorectal Cancer Patients Reveal Population-Specific Insights and Variants

**DOI:** 10.64898/2026.11.19.26368287

**Authors:** Zubayr Kader, Naaziyah Abdulla, Brendan Bebington, Ananyo Choudhury, Phelelani Mpangase, Craig Kinnear, Mandeep Kaur

## Abstract

Colorectal cancer (CRC) genomic studies have largely underrepresented African populations, limiting understanding of disease biology in these groups. This study addressed this gap by generating integrated genomic and transcriptomic data from South African (SA) CRC patients, establishing a foundation for future cancer studies.

Bulk RNA sequencing of 32 samples identified several differentially expressed genes between tumour and normal samples, with *AGAP3*, *AMPD2*, *ANKRD10*, and *SALL4* showing strong diagnostic performance (average sensitivity 99.9%, specificity 94.22%, PPV 99.9%, NPV 89.5%). *SALL4* performed best and was validated using TCGA data and Kaplan-Meier survival analysis. Consensus Molecular Subtype classification revealed a distinct distribution, with enrichment of metabolic (CMS3, 30%) and mesenchymal (CMS4, 30%) subtypes and under-representation of canonical CMS2 (21%). *APC* mutation rates were lower than in European patients (∼55% vs ∼75%). Functional analysis identified inflammatory pathways as primary drivers of carcinogenesis, with viral influences exacerbating the inflammatory tumour microenvironment. Altered neural pathways in tumour samples suggested a consequential gut-brain axis loop as cancer infiltrates enervated tissue, an increasingly relevant field to which these data contribute.

Whole-genome sequencing of 12 samples revealed pathogenic and likely pathogenic variants with transcriptomic differential expression; *SMPD1*, *PERM1*, *PRRT2*, and *RP1* emerged as potential multi-omics markers, while novel mutations (*KDMcA*, *C2CD2*, *TAF2*, *SLCcA1c*, *OR5M10*, *PITRM1*, *TRIMc4B*, *CARD11*) were identified as previously unrecognised contributors to CRC progression in African samples. These findings expand genomic resources for African cancer research and provide new insights into the molecular landscape and tumourigenesis mechanisms of CRC in SA patients, supporting the development of population-specific precision oncology approaches.

## INTRODUCTION

Colorectal cancer (CRC) continues to exhibit an increasing rate of incidence across the globe, with mortality rates predicted to exceed 70% in low and middle income countries [1]. In South Africa (SA), GLOBOCAN estimates a 78% increase in cancer incidence by 2030, which is largely attributed to reduced access to healthcare where many patients are unable to afford screening programmes [2]. This often results in late-stage presentation with poor patient prognosis. The development of population-specific early screening programmes require availability of genetic data from African patients, which is currently non-existent or extremely limited. This critical gap can be addressed through next generation sequencing (NGS) data generation and its analysis, however, individuals of African descent represent less than 3% of the global genomic data available for cancer research [3]. Additionally, public genomic datasets have been overpopulated with data from the USA, Europe and Asia, with research that is not easily extrapolated to the local population and health system of developing countries [4, 5]. In the United Kingdom, the 100,000 Genome Project comprehensively profiled the genomic landscape of 2,023 CRC cases across England, providing a robust foundation for population level genomic analyses. Complementary efforts in Korea employed RNA sequencing to identify differentially expressed genes in CRC patients, while single cell sequencing studies in China have offered detailed insights into the genomic and transcriptomic heterogeneity underpinning the disease [6–9]. A study conducted in the USA utilised genomic sequencing of patient cohorts with African and European ancestry, revealing significant disparities in CRC incidence and mortality between these groups relative to other ancestral populations [10]. These findings point to a distinct carcinogenic mechanism and potential targeted treatment strategies. Interestingly, patients with African ancestry have been associated with a distinct genomic landscape, characterised by higher odds of *APC*, *KRAS* and *PIK3CA* mutations, yet a lower incidence of *BRAF* mutations [11] in contrast to European populations, where *BRAF* mutations are more prevalent [10]. Furthermore, microsatellite instability (MSI) status is consistently reported at a higher level in African patients compared to Europeans, suggesting a unique mechanism of carcinogenesis when compared to traditional pathways such as Wnt [12]. While whole exome sequencing has been performed on sub-Saharan African patients [13], no comprehensive genomic and transcriptomic study has been performed on CRC patient tissue data obtained from a SA patient cohort.

Importantly, SA represents a uniquely diverse population, shaped by a complex mixture of ancestries, environmental exposures, and lifestyle factors. This makes it a significant setting for uncovering novel genetic and molecular features of CRC that may differ from those observed in other regions. To date, there is no set of NGS data comprising RNA-sequencing (RNA-seq) and whole genome sequencing (WGS) from a singular African country. Therefore, in this seminal study, local SA CRC patient tissue samples were collected, sequenced using RNA-Seq and WGS and analysed with bioinformatics tools to produce novel genomic data, elucidating the molecular mechanisms and potential population-specific cancer biomarkers.

## METHODS

### Patient recruitment

The study included sample collection from the Wits Donald Gordon Medical Centre, Park Town, Johannesburg, SA under the Wits Human Research Ethics Committee (Medical) clearance (No: M210233). The inclusion criteria for this study were SA individuals between the ages of 18–85 diagnosed with colon or rectal cancer. All 32 patients underwent informed consent for the collection, processing and storage of their tissue samples. All research was performed with relevant guidelines and regulations to ensure patient anonymity and protection of personal data.

### Collection of Local Tissue Samples and Nucleic Acid Isolation

Tissue specimens were isolated from the resected tumours as well as at a distant site 10–15 cm away from the tumour periphery termed “normal tissue”. The tissues were placed in 50 ml falcon tubes containing Dulbecco’s Modified Eagle Medium (Thermo Fisher Scientific, USA) supplemented with the antibiotic Primocin (Invitrogen, USA), 1x Glutamax (Thermo Fisher Scientific, USA) and 10 mM HEPES (Thermo Fisher Scientific, USA). The tissue samples were washed twice with 1x Dulbecco’s phosphate buffered saline (Sigma Aldrich, UK) supplemented with Primocin (Invivogen, USA), and subsequently stored in RNALater™ (Thermo Fisher Scientific, USA) for nucleic acid isolation.

Total RNA was isolated using the Qiagen RNeasy Mini Kit (Qiagen, Germany) and total genomic DNA was isolated using the Qiagen DNeasy Blood C Tissue Kit (Qiagen, Germany) according to the manufacturer’s protocols. Nucleic acid purity and integrity was validated prior to downstream processing using the Qubit Fluorometer (Thermo Fisher Scientific, USA).

### Library Preparation and NGS Processing

Library preparation and sequencing was performed at the South African Medical Research Council Genomics Platform, Cape Town, South Africa. For RNA sequencing of 32 RNA samples, further depletion of rRNA was performed using the MGIEasy rRNA Depletion Kit (MGI, GD, China) according to the manufacturer’s protocol. Additional globin depletion was performed using the NEBNext Globin C rRNA Depletion Kit (Human) (New England Biolabs, MA, USA) according to the manufacturer’s protocol. The sequencing library was prepared using the MGIEasy RNA Library Prep Set (MGI, GD, China) according to the manufacturer’s protocol. Sequencing was performed in paired end reads of 2×100bp using the DNBSEQ-G400 High-throughput Sequencing Set on the DNB-SEQ-G400 high-throughput sequencer (MGI, GD, China).

For whole genome sequencing (WGS), the sequencing libraries using DNA were prepared for 12 samples (6 normal vs 6 cancer, of which 6 were chemotherapy treated and 6 untreated) using the MGIEasy Universal DNA Library Prep Set (MGI, GD, China) and sequenced in paired end reads of 2×150bp on the DNBSEQ-G400 high-throughput sequencer (MGI, GD, China).

The raw FASTQ sequence files were pre-processed and aligned to the GRCh38.p14 human genome and transcriptome from the Gencode database (https://www.gencodegenes.org/human/). For RNA-Seq, the Salmon tool was used to pseudo-align to the human transcriptome to quantify transcripts and generate count matrices [14].For WGS, samples were aligned to the human genome using BWA-MEM, and variants were called using Google Research DeepVariant and GLNexus [15, 16].

### RNA-Seq Analysis: Differential Expression and Functional Enrichment

Biologically relevant genes and processes were selected using differential expression analysis and functional analysis as described below. Preliminary principal component analysis was performed to visualise the variance in gene expression profiles between the normal and cancer samples. The raw gene counts were normalised using variance stabilising transformation to remove artificial variation between the samples and were then used to calculate the mean count value per gene across the patient samples. The DESeq2 negative binomial model was used to identify differentially expressed (DE) genes [17]. A Wald-test was used to test if a log_2_ fold change between the sample groups is significantly different from zero, with a Benjamini-Hochberg correction to adjust the p-value to control for false positives, where significance was set to p<0.05. Genes with positive log fold changes were considered as upregulated, while those with negative indicted downregulated genes.

In order to elucidate the biological functions of the DE genes, functional analysis was performed using the Gene Ontology (GO) and Kyoto Encyclopaedia of Genes and Genomes (KEGG) databases [18, 19]. Enrichment, also known as over-representation analysis, assesses the likelihood that functional categories within a specific gene list occur at a significantly higher proportion compared to a background set of genes. Data sets comprising the significant DE genes were used as input of primary interest, while all DE genes, regardless of their significance, were used as background data sets. Functional enrichment was performed in R using clusterProfiler as well as the web tool ShinyGO [20, 21].

The solitary examination of individual GO processes and KEGG pathways yields limited biological insights. Network plots serve as a robust visualisation tool, revealing intricate relationships among genes and processes or pathways and were developed using DE genes for each comparison and leveraging the information from the GO and KEGG database using the Pathview network analysis tool [22].

### Whole Genome Sequencing Analysis

The sequences were processed using BWA-mem v0.7 to align to the human reference genome GRCh38.p.14, with Google Research DeepVariant v1.8.0 in conjunction with GLNexus v1.4.1 used to call variants and perform joint-calling [15, 16]. Variant analysis was performed using the Genome Analysis Toolkit [23]. The ’Funcotator’ tool, serving as a functional annotator, was employed for the annotation of variants in the filtered VCF files, utilising the pre-installed somatic data sources for querying reference SNP cluster ID (rsID) and single nucleotide polymorphism database identifier (dbSNP). Further variant analysis was performed using the OpenCRAVAT python package to annotate COSMIC and ClinVar information [24].

The variants were filtered as follows: the coding variants were isolated and assigned to the ClinVar database for clinical information, and the variants present in ClinVar were extracted [25]. Then, this set of variants was mapped to the COSMIC database, and the present genes were used for biological interpretation [26].

In order to create a holistic view of the genome-to-transcriptome, variants of interest were identified and isolated from the significant DE gene RNA-seq dataset to analyse their expression profiles.

### Biomarker Identification and in-silico Validation

To evaluate the predictive prognostic potential of the selected significantly DE genes (p<0.05 and log_2_FC > 1 or log_2_FC < 1), their expression was examined with respect to the sensitivity, specificity and precision performance measures [27]. These evaluations employ parameters to determine whether a patient expressed the gene of interest. The following tests were conducted using an R script to establish the parameters: TP, representing true positives or patients who have CRC and express the gene; TN, denoting true negatives or patients who do not have CRC and do not express the gene; FP, indicating false positives or patients who do not have CRC but express the gene; and FN, representing false negatives or patients who have CRC but do not express the gene.

Sensitivity and specificity reflect the ability of a gene to correctly classify disease and non-disease samples, respectively. Positive Predictive Value (PPV) indicates the probability that a sample predicted to be positive truly represents disease, while Negative Predictive Value (NPV) indicates the probability that a sample predicted to be negative is truly disease-free.

To evaluate expression levels in CRC, the combined colon cancer database integrated into Kaplan-Meier Plotter (https://www.kmplot.com/analysis/) was mined, which includes data from 2,137 tumour samples from 17 independent cohorts based on Affymetrix Human Genome U133A Arrays [28]. This produced Kaplan-Meier survival analysis plots for each biomarker, as well as the gene signature comprising of all the biomarkers. Additional validation was performed through box plot expression analysis using GEPIA3 (https://gepia3.bioinfoliu.com/) and the TCGA COAD and READ datasets [29]. Further review of potential biomarkers was done by assessing their presence in the CoReCG Database [30].

## RESULTS

### Patient Sample Details

Table 1 details the age, gender, ancestry, diagnosis and chemotherapy status of 32 tissue samples from 21 patients, where 8 were females and 13 were males. The age of patients ranged from 38 to 83 years. Out of 32 samples, 16 were chemotherapy treated and 16 were chemotherapy naïve (herewith noted as chemotherapy treated and untreated respectively). Stages of tumours were available for some patients, with seven in Stage 1, eight in Stage II, six in Stage III and four in Stage IV. There were 18 cancer and 14 normal samples. Tumour stage and MSI status varied from early to late stages, with both MSS and MSI tumours. Historical data for some patients was not accessible.

**Table 1:** Basal Characteristics and demographics of Enrolled Patients.

| Sample Name | Age Range (years) | Gender | Ancestry | Diagnosis | Chemotherapy | Site | Tumour Stage | MSI/MSS Status |
|---|---|---|---|---|---|---|---|---|
| B04N | 51-55 | F | Indian | Normal | Yes | Colon | IV | -- |
| B05N | 56-60 | M | White | Normal | Yes | Rectal | IIA | MSS |
| B06N | 61-65 | M | White | Normal | No | Colon | IIB | -- |
| B07C | 81-85 | M | White | Cancer | No | Colon | IIIB | MSS |
| B07N | 81-85 | M | White | Normal | No | Colon | IIIB | MSS |
| B08C | 81-85 | F | White | Cancer | No | Colon | IIA | -- |
| B09C | 61-65 | F | White | Cancer | No | Colon | - | -- |
| B09N | 61-65 | F | White | Normal | No | Colon | - | -- |
| B10C | 51-55 | M | Indian | Cancer | Yes | Colon | IIIA | -- |
| B10N | 51-55 | M | Indian | Normal | Yes | Colon | IIIA | -- |
| B11C | 61-65 | M | White | Cancer | Yes | Colon | IIA | MSI |
| B11N | 61-65 | M | White | Normal | Yes | Colon | IIA | MSI |
| B12C | 61-65 | M | White | Cancer | Yes | Rectal | IV | MSS |
| B13C | 51-55 | M | Black | Cancer | Yes | Rectal | IIIC | -- |
| B14C | 71-75 | F | White | Cancer | No | Colon | I | MSI |
| B14N | 71-75 | F | White | Normal | No | Colon | I | MSI |
| B15C | 56-60 | F | Mixed* | Cancer | Yes | Rectal | IV | MSS |
| B15N | 56-60 | F | Mixed* | Normal | Yes | Rectal | IV | MSS |
| B16C | 71-75 | M | White | Cancer | No | Colon | - | -- |
| B17C | 66-70 | F | Indian | Cancer | No | Colon | I | -- |
| B17N | 66-70 | F | Indian | Normal | No | Colon | I | -- |
| B18C | 56-60 | M | White | Cancer | Yes | Rectal | I | MSS |
| B18N | 56-60 | M | White | Normal | Yes | Rectal | I | MSS |
| B19C | 71-75 | M | White | Cancer | Yes | Colon | - | -- |
| B19N | 71-75 | M | White | Normal | Yes | Colon | - | -- |
| B20C | 71-75 | M | White | Cancer | No | Colon | IIB | -- |
| B21C | 61-65 | M | Indian | Cancer | Yes | Colorectal | I | -- |
| B22C | 61-65 | M | White | Cancer | Yes | Rectal | IIIB | -- |
| B26C | 36-40 | F | White | Cancer | No | Rectal | - | -- |
| B26N | 36-40 | F | White | Normal | No | Rectal | - | -- |
| B27C | 51-55 | F | White | Cancer | No | Colon | IIA | MSI |
| B27N | 51-55 | F | White | Normal | No | Colon | IIA | MSI |

*\*Individuals with mixed ancestry*

### Identification of candidate biomarkers gene signature facilitating carcinogenesis

DE analysis of RNA-Seq data comparing tumour and matched normal samples enabled the systematic identification of significantly up- and downregulated genes, providing insight into the transcriptional alterations associated with carcinogenesis and supporting downstream functional and pathway-level analyses. This analysis also led to the identification of candidate biomarker genes for testing. The sequences passed quality checks (see Supplementary Figures S1 C S2).

The DE analysis was performed in three contrasts: Cancer vs Normal (Contrast 1: all data), Cancer vs Normal (Contrast 2: untreated), Cancer vs Normal (Contrast 3: treated). The results in the Figures presented in the main text focus on Contrast 1 (cancer vs normal – all data), with further results for Contrasts 2 and 3 attached as Supplementary information.

The principal component analysis plot in Figure 1A shows slight separation between the normal and cancer clusters possibly because normal tissues are derived from the cancer patients but still variance is visible between cancer and normal tissues. The groups also show intra-cohort variance. There was no visible clustering according to microsatellite status or stage.

**Figure 1:**
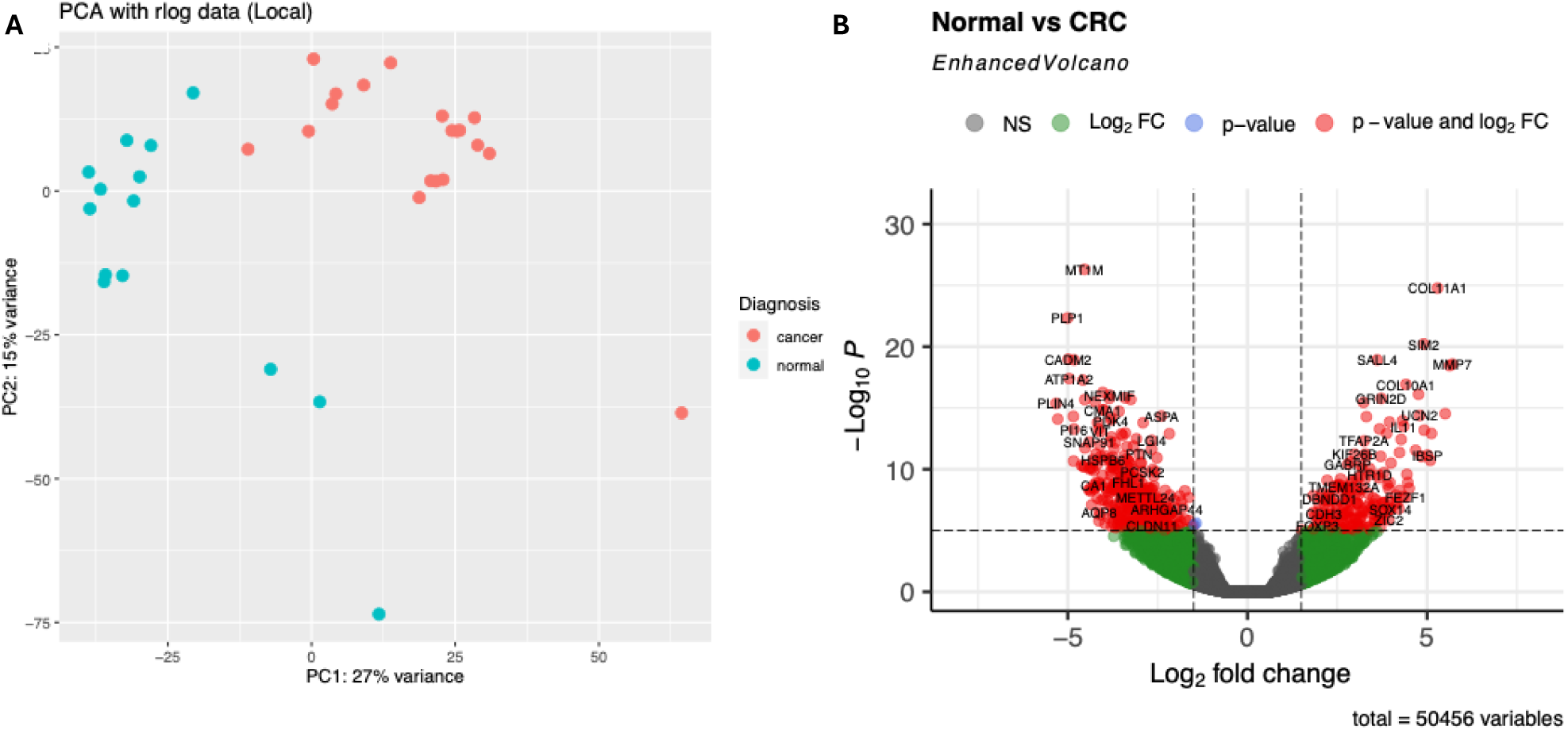
(A) Principal component analysis plot showing variation between the cancer and normal gene expression profiles. (B) Volcano plot visualising the differentially expressed genes across the patient cohort. the X-axis showing the log_2_FC, and the Y-axis showing the log_10_ P-Value. The horizontal and vertical dashed lines signify the area of genes that satisfy both thresholds, with the red dots indicating genes significantly DE (genes that meet the p<0.05 and log_2_FC>0.58 thresholds). The thresholds of p< 0.05 and log_2_FC 1.5 were used to identify significantly differentially expressed genes (red dots).

The volcano plot in Figure 1B shows the spread of DE genes between cancer vs normal. The negative binomial model from DESeq2 was used in R to identify DE genes. There were 582 significantly (p<0.05) upregulated genes, and 1113 significantly (P<0.05) downregulated genes (see Supplementary Excel WS1 and WS2). In treated samples, there appeared to be higher significance for upregulation of genes, while in untreated samples for downregulation (see Supplementary Figures S3 C S4). The individual genes that were most significantly upregulated and downregulated according to their adjusted p-values are displayed in Table 2 C Table 3, respectively.

**Table 2:** The Top 10 upregulated genes with brief description and Log2 fold change values (shaded according to increasing value from green to red)

| Gene Symbol | Description | Log<sub>2</sub> Fold Change | *p*-value (adjusted) |
|---|---|---|---|
| **COL11A1** | Type XI collagen alpha unit [31] | 5.88 | 1.46e-27 |
| **SERPINB3** | Modulates host immune against tumour cells [32] | 25.59 | 1.26e-25 |
| **SALL4** | Transcription factor with a key role in the maintenance and self-renewal of embryonic and hematopoietic stem cells [33] | 3.80 | 2.80e-23 |
| **SIM2** | Transcription factor that may be a master gene of CNS development in cooperation with Aryl Hydrocarbon Receptor Nuclear Translocator [34] | 5.47 | 1.34e-22 |
| **CXCL8** | May promote cell proliferation and EMT [35] | 6.83 | 9.49e-21 |
| **MMP7** | Encodes matrix metalloprotease-7 [36] | 6.99 | 9.49e-21 |
| **PLEKHN1** | Involved in the hypoxia induced apoptotic pathway [37] | 3.39 | 9.16e-20 |
| **COL10A1** | Type X collagen alpha unit [38] | 4.88 | 1.31e-19 |
| **GRIN2D** | Glutamate receptor gene involved in the RAD/MAP kinase cascade [39] | 3.99 | 3.06e-19 |
| **SPP1** | Involved in the cytokine activity and ECM binding [40] | 5.45 | 2.00e-18 |

\*\*\* Genes are presented in descending order of adjusted p-values

**Table 3:** The Top 10 downregulated with brief descriptions and Log2 fold change values (shaded according to increasing value from green to red)

| **Gene Symbol** | **Details** | **Log<sub>2</sub> Fold Change** | ***p*-value (adjusted)** |
|---|---|---|---|
| **MT1M** | Encodes metallothioneins and may act as a tumour suppressor [41] | 4.83 | 2.42e-30 |
| **PLP1** | The major myselin protein in the CNS [42] | 5.52 | 7.68e-25 |
| **ASPA** | Encodes aspartoacylase, may be a tumour suppressor in prostate cancer [43] | 2.45 | 2.59e-21 |
| **PRIMA1** | Encodes enzyme binding and protein-membrane adaptor activity, may have an antitumour effect [44] | 5.38 | 4.37e-21 |
| **CADM2** | Encodes cell adhesion molecule 2, acts as a tumour suppressor [45] | 5.64 | 7.49e-21 |
| **SCN9A** | Involved in sodium channels related to the nervous system [46] | 2.22 | 1.71e-20 |
| **ABCA8** | Encodes a protein involved in the ATP-binding cassette and cholesterol regulation [47] | 3.41 | 4.51e-20 |
| **RIMS4** | Involved in transmembrane transporter binding [48] | 5.12 | 1.86e-19 |
| **ATP1A2** | Encodes for an ATPase subunit, abnormally expressed in breast cancer [49] | 5.65 | 2.45e-19 |
| **FXYD1** | Encodes phospholemman, involved in ATPase subunit [50] | 4.14 | 5.74e-19 |

\*\*\* Genes are presented in descending order of adjusted p-values

Overlap analysis of DE genes revealed a substantial shared transcriptional signature between chemotherapy treated and untreated tissue samples (Supplementary Figure S5). Of all DE genes, 1,497 were common to both conditions, while 1,645 and 1,005 were unique to treated and untreated samples, respectively, indicating treatment-associated modulation of an underlying disease-driven expression program.

When stratified, upregulated genes showed limited overlap (438 shared genes), with substantial condition-specific upregulation in treated (580 genes) and untreated (455 genes) samples, suggesting that treatment preferentially alters gene activation. In contrast, downregulated genes exhibited strong overlap (1,059 shared genes), with a larger number uniquely downregulated in treated samples (1,066 vs 551), consistent with conserved repression of tumour-associated genes and additional treatment-associated downregulation.

Overall, treatment modifies rather than replaces the core transcriptional landscape, with more pronounced effects on upregulated genes.

### Functional Analysis of differentially expressed reveal altered organ function and enrichment of inflammatory pathways as a key driver of carcinogenesis

The functional enrichment of the DE genes using GO and KEGG resulted in both individual processes and pathways, as well as an informative network view. The top 20 GO processes and KEGG pathways for the up and downregulated genes can be viewed in Supplementary Figures S10 and S11. The network plots on the interaction of the processes and pathways are shown in Figure 2, while the networks for the treated and untreated contrasts are shown in Supplementary Figures S6 to S9. Upregulated processes include those related to DNA replication, cell cycle and chromosomes suggesting genetic changes within the cancer tissue, whereas downregulated changes involve those related to respiration and energy possibly a by-product of the change in tumour microenvironment (TME). In terms of pathways, upregulated pathways involve inflammatory related signalling in IL-17, cytokine interactions, p53 and TNF pathways. Interestingly, there is an upregulated pathway relating to viral interactions, further characterising the TME as inflamed. Downregulated pathways primarily consist of normal physiological processes in muscle contraction and circulatory system that may have been disrupted by the cancer progression in the organ. Previous studies across multiple cancer types have identified gene expression signatures of carcinogenesis involving genes that also regulate normal cellular functions, including in bladder and lung cancers [51].

**Figure 2:**
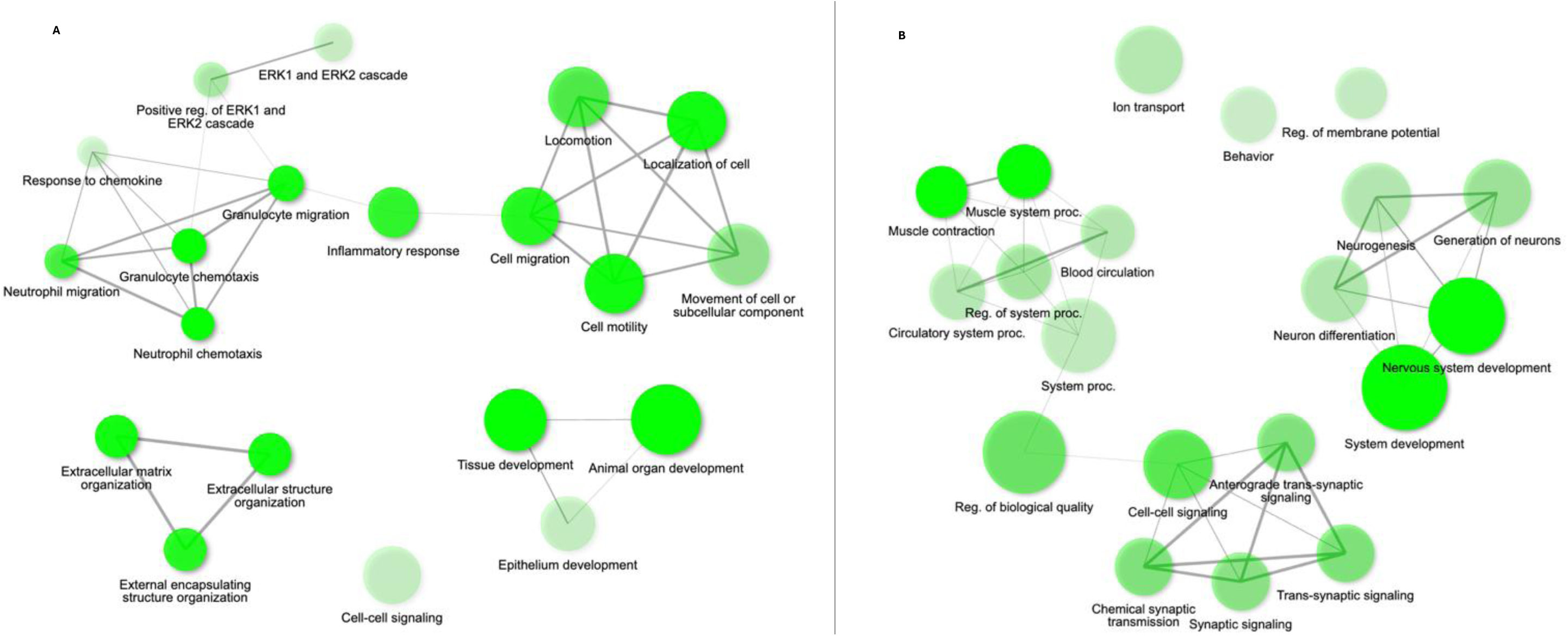
Network plot for the Gene Ontology (GO) Biological Processes for the (A) up- and (B) downregulated genes from the RNA-Seq analysis. False discovery rate was set to 0.05, with the minimum pathway size set to 2 and the top 20 pathways as ordered by p-value are shown.

### Chemotherapy treatment is associated with metabolic reprogramming and enhanced immune activation

To assess the impact of chemotherapy on tumour biology, DE analysis compared chemotherapy treated and untreated samples. This comparative approach enabled the identification of transcriptional changes associated with treatment exposure, providing insight into treatment-related modulation of carcinogenic, metabolic, and immune-related processes.

A subset of the results is displayed in Figure 3 for each contrast, namely Treated Normal vs Treated Cancer, and Untreated Normal vs Untreated Cancer.

**Figure 3:**
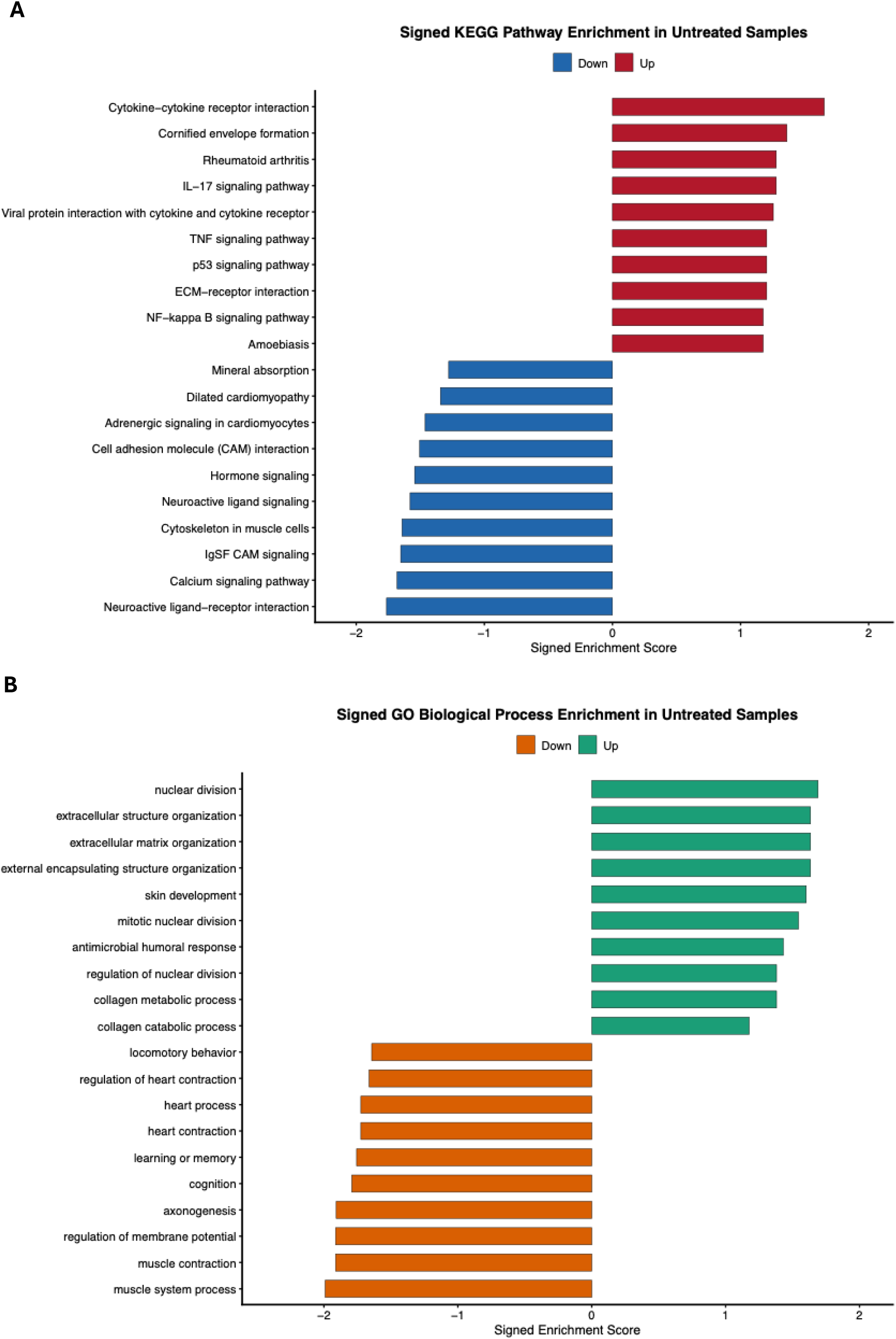
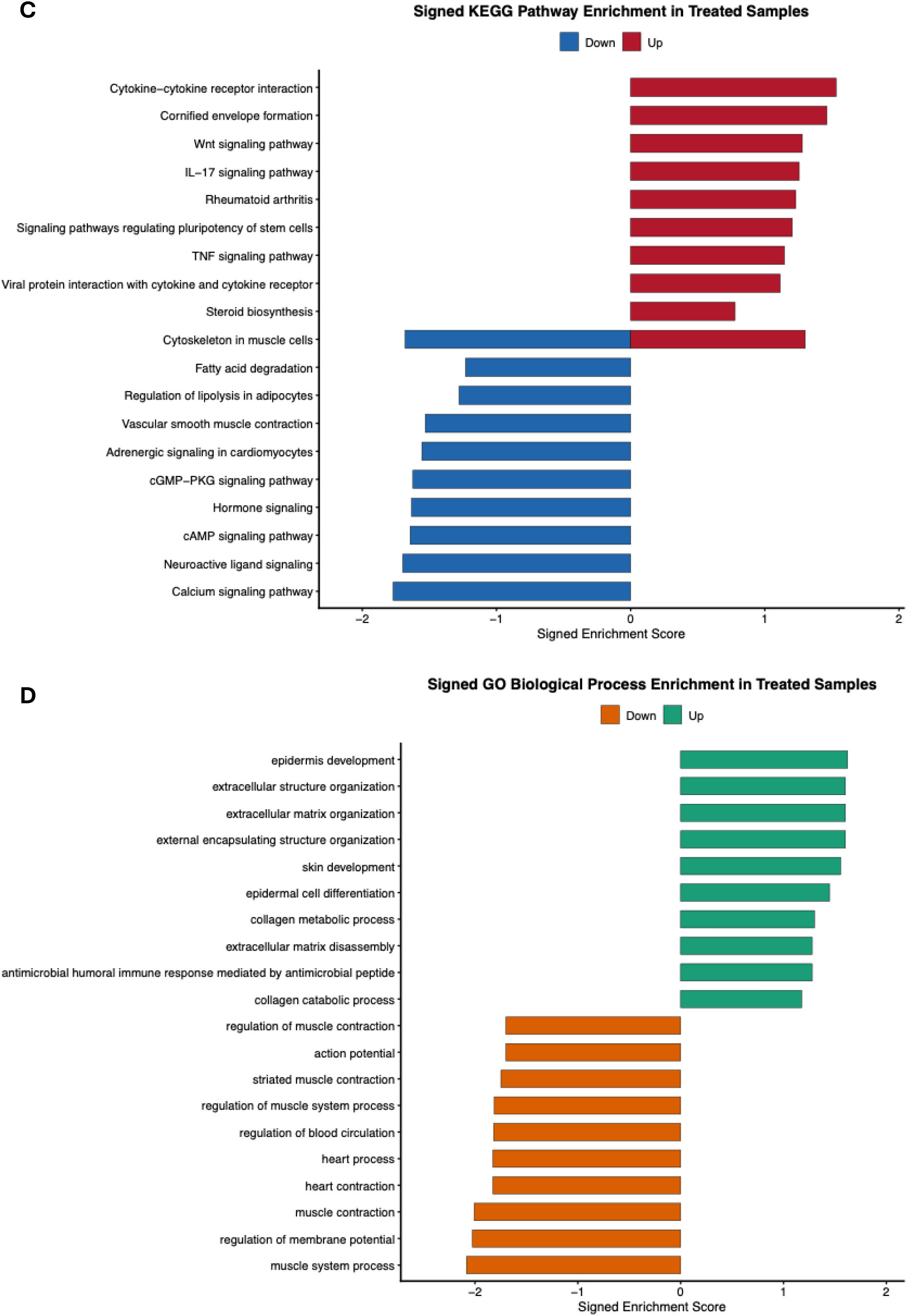
Enriched KEGG Pathway and GO Biological Processes for the different treated and untreated sample groups. (A) KEGG Pathway Enrichment Plot for untreated samples (n = 1c). (B) GO Biological process Enrichment Plot for untreated samples (n = 16). (C) KEGG Pathway Enrichment Plot for treated samples (n = 16). (D) GO Biological process Enrichment Plot for untreated samples (n = 16). All figures were created with DE genes p<0.05.

In untreated samples, Figure 3A suggests a remodelling phenotype is present with dysregulated normal function of the organ such as muscle contraction and membrane potential. Figure 3B further shows there is strong upregulation of inflammatory, and remodelling pathways with a downregulation in homeostatic signalling pathways, namely calcium signalling and hormone signalling.

Chemotherapy treated samples in Figure 3C show a strong remodelling phenotype present, and additional downregulation of processes related to normal organ function in muscle contraction and blood circulation. Treated samples appear to have a stronger inflammatory signalling upregulation through various pathways (Figure 3D) such as IL-17 signalling pathway, TNF signalling and cytokine-cytokine pathways, with a more prevalent dysregulation of homeostatic functions as compared to the untreated samples.

Taken together, the data indicate that while untreated tumours already display classic hallmarks of cancer such as metabolic rewiring and stromal remodelling, chemotherapy profoundly reshapes tumour biology. After treatment, cancer tissue relative to normal is dominated by inflammatory signalling, immune activation, cellular reprogramming, and aggressive tissue remodelling programmes. This suggests that chemotherapy not only disrupts tumour cells but also drives the tumour ecosystem into a high-stress, regenerative and adaptive state, in which surviving cells engage with pathways associated with plasticity, invasion, and potential therapy resistance.

#### Consensus Molecular Subtypes

Consensus Molecular Subtype (CMS) classification using CMS classifier [52] revealed a distinct subtype distribution in the SA CRC cohort, with enrichment of the metabolic (CMS3, 30%) and mesenchymal (CMS4, 30%) subtypes, and a relative under-representation of the canonical CMS2 subtype (21%).

### Whole-genome sequencing identifies novel and functionally relevant variants linked to transcriptomic and pathway-level alterations

Whole-genome sequencing was conducted to characterise the variant landscape within the cohort and to identify novel and variants of potential interest. Identified variants were examined alongside transcriptomic and functional enrichment results to explore concordance between genomic variation, gene expression patterns, and implicated biological pathways.

Table 4 displays the patient characteristics of the 12 sequenced samples used for sequencing. Ages ranged from 34 to 70 years old, with four tissue samples from African patients and the remainder from White patients. There was one Stage I sample, five Stage II samples, four Stage III samples and two Stage IV samples. Six samples were chemotherapy treated and 6 were chemotherapy naïve. Majority of the samples were classified as MSS status.

**Table 4:**
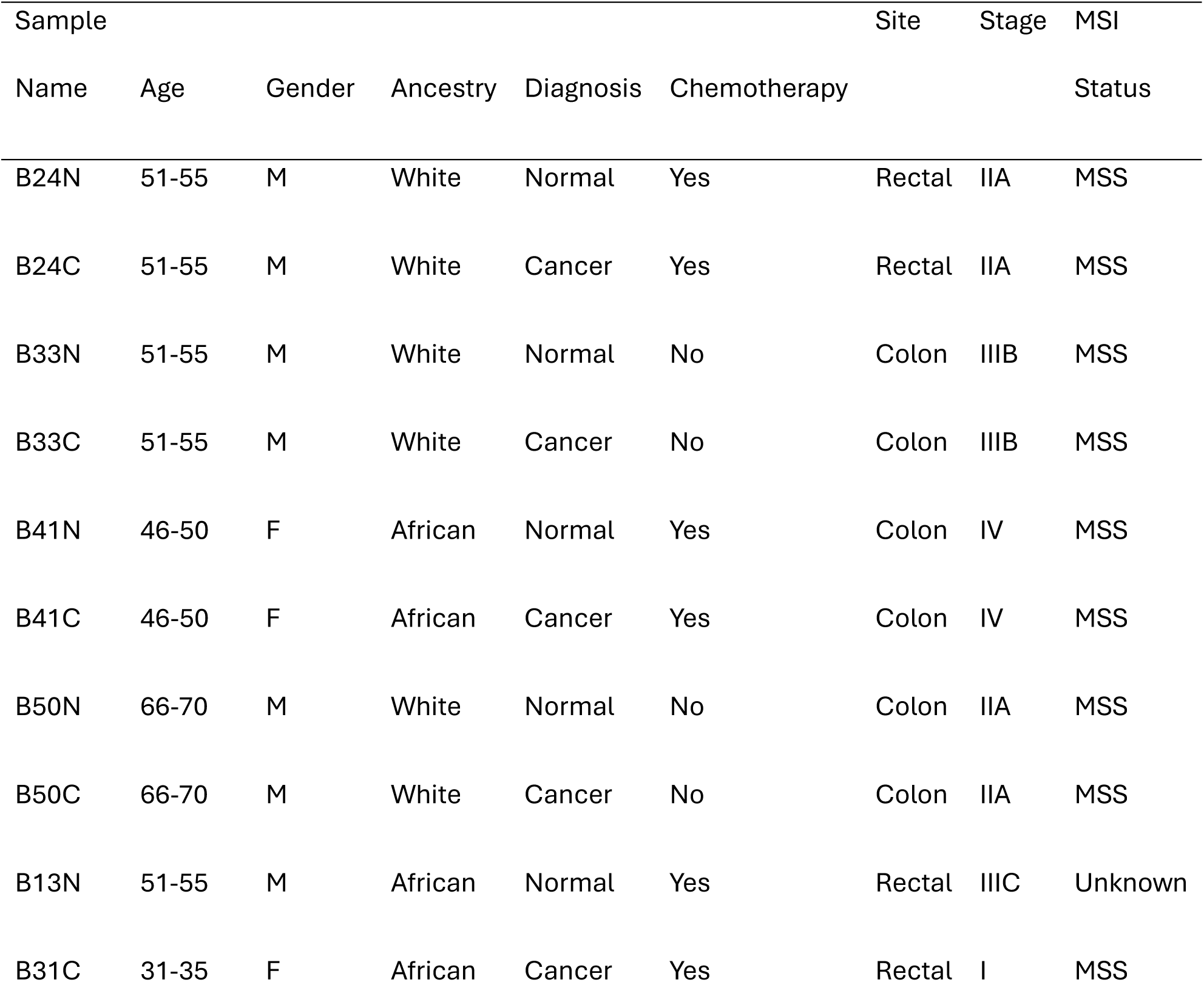
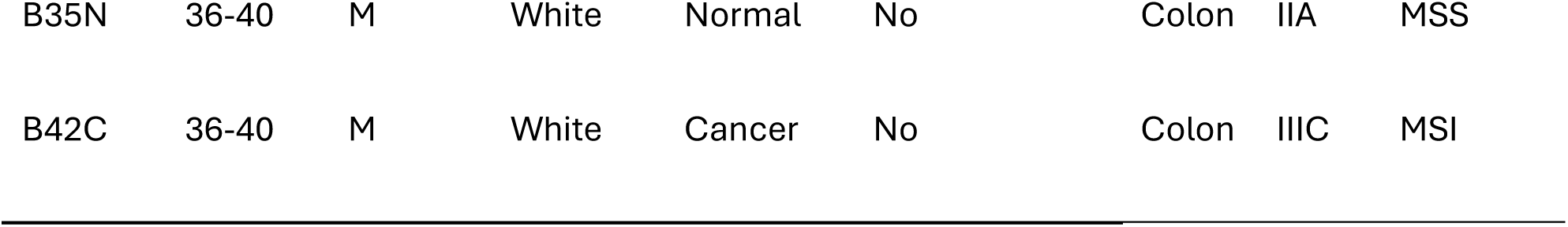
Patient Clinical Characteristics.

The cohort-level joint called VCF file containing all the samples was used for analysis. The total number of unique genetic variants identified was 13,217,568 of which 64,455 were coding variants. Most of the variants were synonymous and missense. Filtering the variants of interest involved annotation to the ClinVar database [25] and the COSMIC database [53]. Of the coding variants, 23,026 (36%) were reported in ClinVar with 14,356 (22%) appearing in COSMIC. This initial collection of variants was used for the downstream analysis to understand the variant landscape in SA CRC. Supplementary Table S1 shows the different variant types present in the main coding regions.

While filtering, particular focus was paid to the known driver gene *APC* with prior research indicating variation in *APC* mutations between different groups of differing ancestry [54]. Analyses yielded 16 variants of interest shown in Table 5.

**Table 5:**
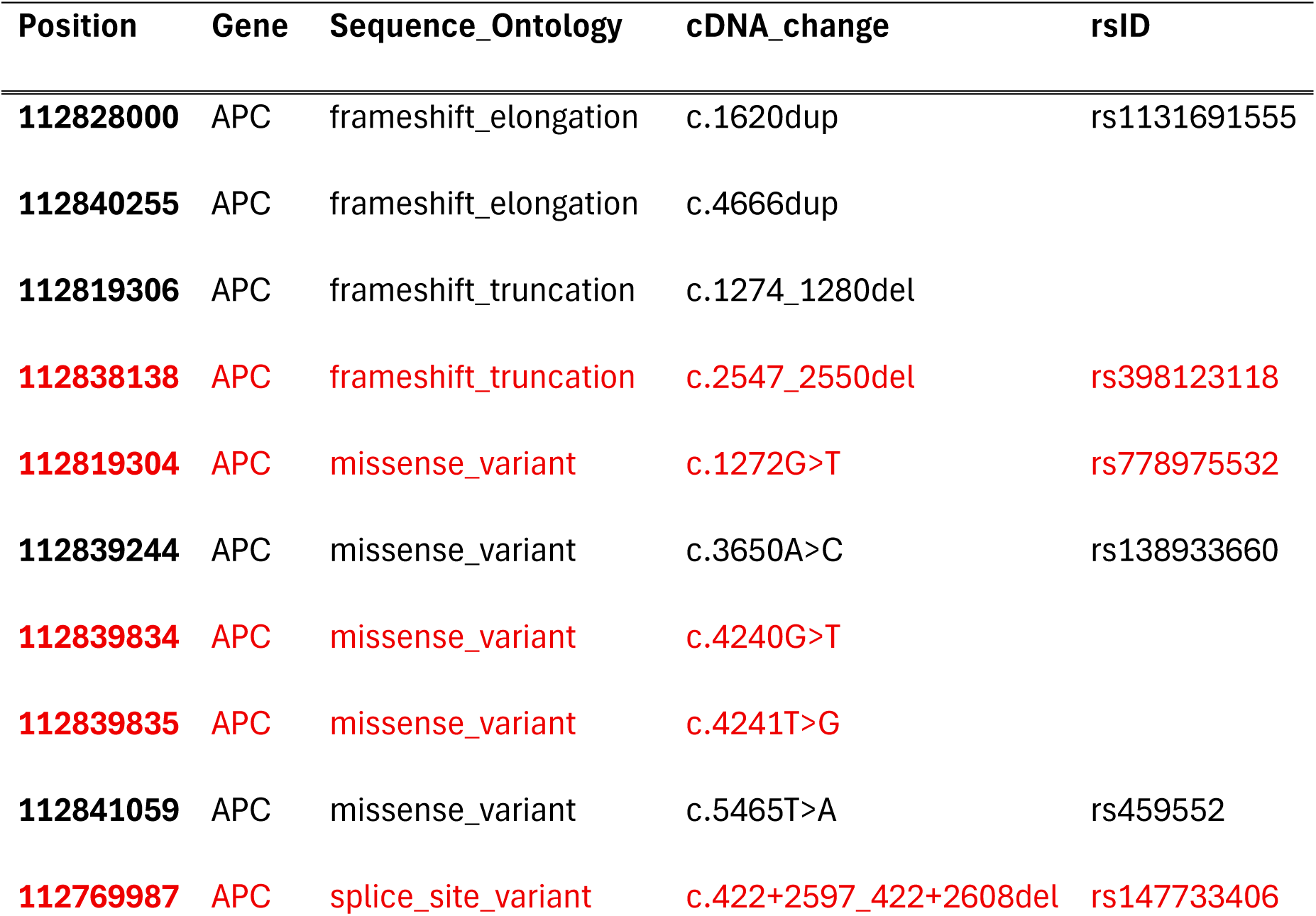
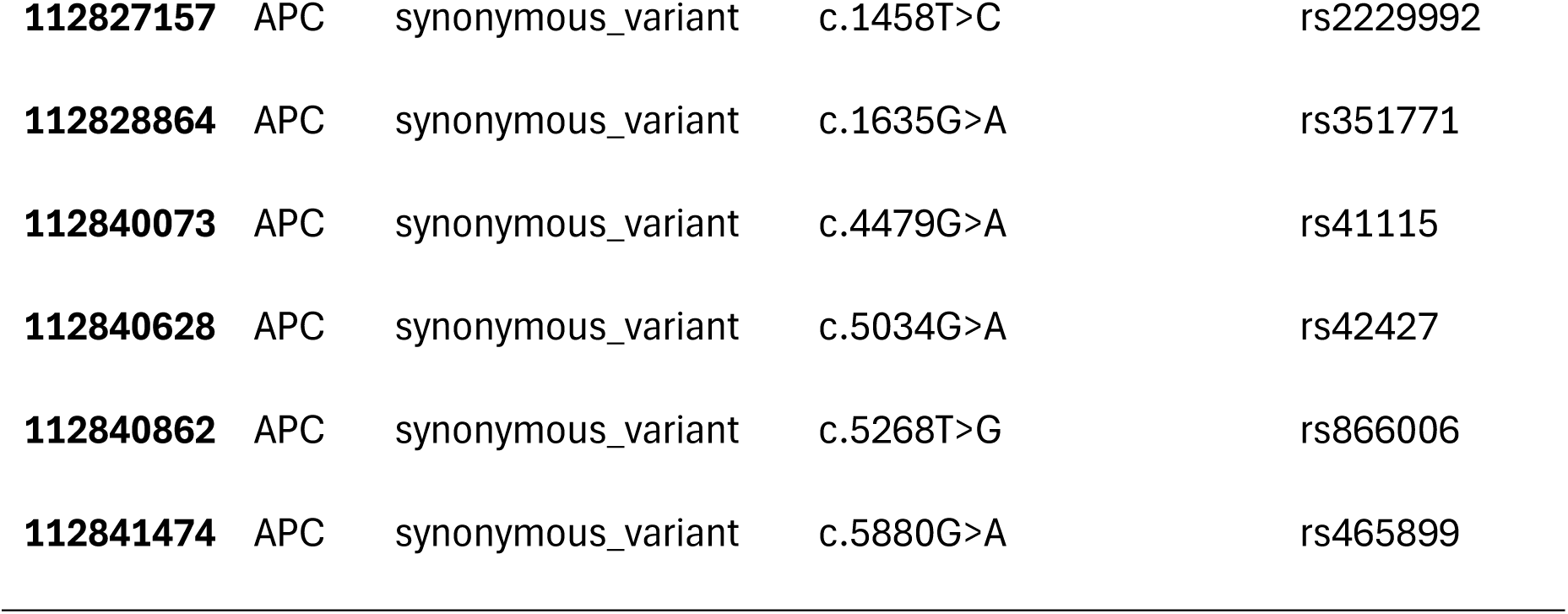
Variants present in the APC gene amongst the sample cohort.

Five of the above variants are absent from both COSMIC and ClinVar (in red text colour), supporting the hypothesis that uncommon *APC* variants may contribute to disease severity in African populations. Tumour sequencing revealed recurrent inactivating mutations in *APC*, consistent with the canonical Wnt pathway driven tumorigenesis. Multiple independent truncating variants were observed, including frameshift duplications and deletions (c.1620dup, p.Gln541ThrfsTer19; c.4666dup, p.Thr1556AsnfsTer3; c.1274_1280del, p.Glu425ValfsTer27; c.2547_2550del, p.Asp849GlufsTer11), as well as a splice-site deletion (c.422+2597_422+2608del), all predicted to result in premature protein truncation and loss of function.

The original 64,455 variants were filtered to isolate somatic variants (variants present in only the cancer samples), which resulted in 8427 variants of interest. There were eight functional previously unrecorded variants present in the cancer samples only, and are displayed in Table 6:

**Table 6:** Functional genetic variants exclusively present in cancer samples.

| **Gene** | **Sequence Ontology** | **cDNA Change** | **Description** |
|---|---|---|---|
| *KDM6A* | nonframeshift deletion | c.312+14_312+5299del | Regulates cell differentiation [55] |
| *C2CD2* | frameshift deletion | c.954-480_953+1del | Potential biomarker for CRC [56] |
| *TAF2* | nonframeshift deletion | c.300-3600_299+1del | Regulates essential cell growth proteins [57] |
| *SLC6A16* | frameshift deletion | c.1230-5448_1229+2del | Involved in neurotransmitters, possible biomarker for lung cancer [58] |
| *OR5M1* | frameshift deletion | c.429_432del | Olfactory receptor downregulated in breast cancer [59] |
| *PITRM1* | stopgain | c.271_272ins | Mitochondrial enzyme, linked with neurodegenerative disorders [60] |
| *TRIM64B* | frameshift deletion | c.377del | Regulates immune response, identified cancer driver [61] |
| *CARD11* | frameshift insertion | c.1663dup | Regulates CD8 tumouricidal function [62] |

The 14,356 filtered variants that were present in COSMIC involved 4876 individual genes, of which 4834 were mapped to the KEGG database and the pathways are displayed in Figure 4. Many of the genes were involved in the extracellular matrix (ECM) function of the tumour, as well as pathways relating to cancer, metabolism and signalling pathways suggestive of a change in the TME as the tumour progresses. There is further activation of core oncogenic survival pathways in PI3K-Akt and HIF-1 signalling, metabolic reprogramming and pro-metastatic platelet and coagulation signalling.

**Figure 4:**
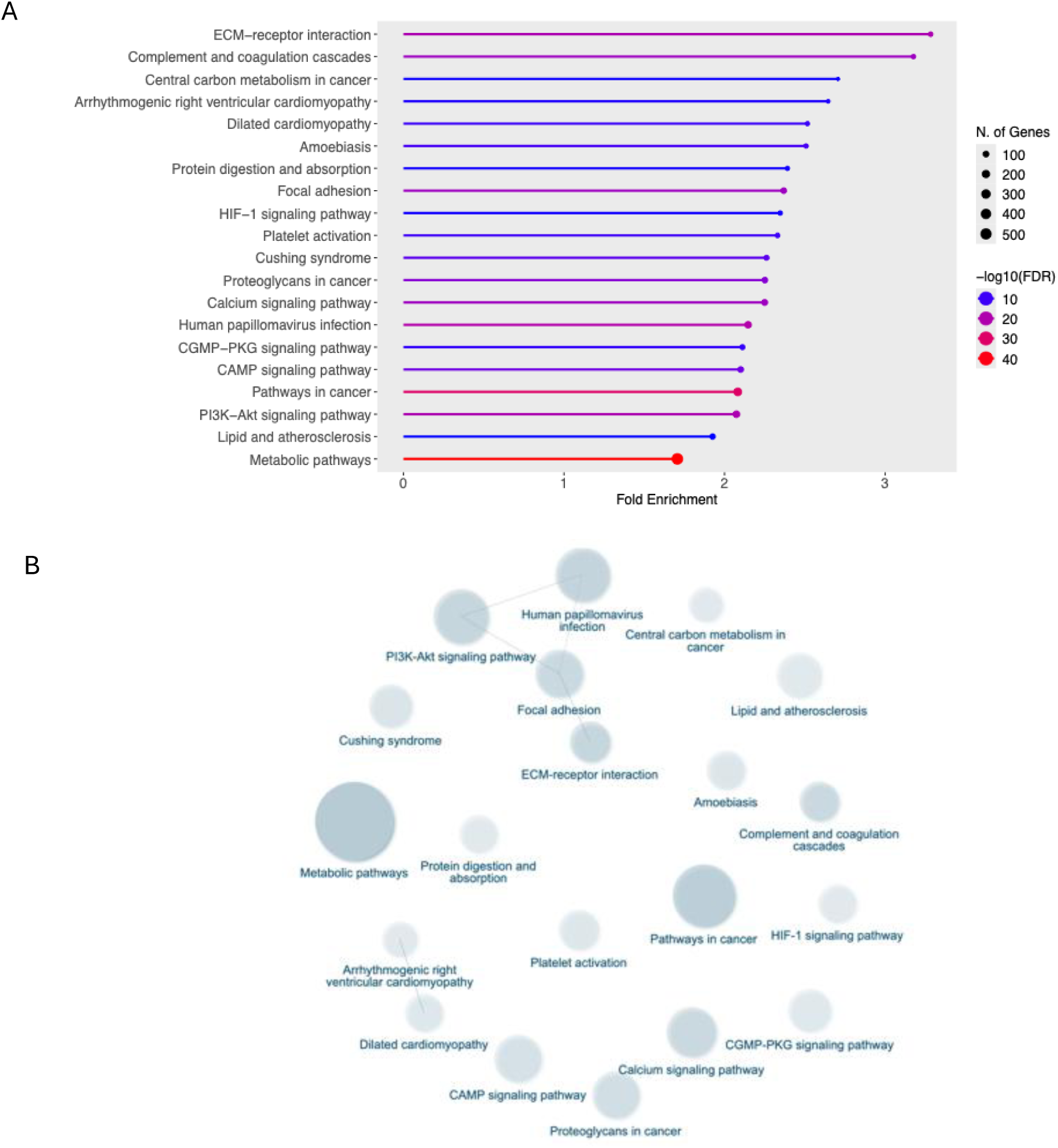
Functional enrichment of variants of interest with genes mapped to KEGG pathways. (A) Shows the bar plot of mapped KEGG pathways. (B) Network interaction plot of the pathways. In (A) the x-axis represents the fold enrichment of the KEGG Pathway terms while the y-axis lists the corresponding pathways. Line colours indicate significance based on the false discovery rate (FDR), ranging from blue (low significance) to red (high significance). Circular endpoints on each line denote the number of genes within each pathway that were present in the gene expression dataset.

In order to further prioritise the variant list, the “pathogenic” and “likely pathogenic” filter was applied to the ClinVar data, resulting in 24 known variants of interest. The full table of variants can be found attached as Supplementary Excel WS3. These variants were then identified in the analysed RNA-Seq data, producing the first genomic and transcriptomic analysis of its kind using South African data. Of the 24 genes with variants, 4 were identified in the transcriptomic expression profile data, *PPARGC1* (Peroxisome proliferator-activated receptor gamma coactivator 1-alpha) And ESRR (estrogen related receptor) Induced Regulator in Muscle 1 (*PERM1)* and RP1 Axonemal Microtubule Associated gene (*RP1)* were upregulated, while Proline Rich Transmembrane Protein 2 (*PRRT2)* and Sphingomyelin Phosphodiesterase 1 (*SMPD1)* were downregulated. Their expression profiles are shown in Figure 5. The upregulation of *PERM1*, a regulator of mitochondrial and oxidative metabolism, suggests increased mitochondrial activity and metabolic reprogramming within tumour tissue. *RP1*, although classically associated with microtubule organisation in retinal cells, was also upregulated, potentially indicating broader alterations in cytoskeletal organisation and cellular architecture. In contrast, *PRRT2*, a gene implicated in membrane-associated signalling, was downregulated, which may reflect disruption of intracellular communication pathways. Similarly, reduced expression of *SMPD1*, a key regulator of sphingolipid metabolism and apoptotic signalling, is consistent with attenuated cell death pathways and increased cell survival in cancer tissue.

**Figure 5:**
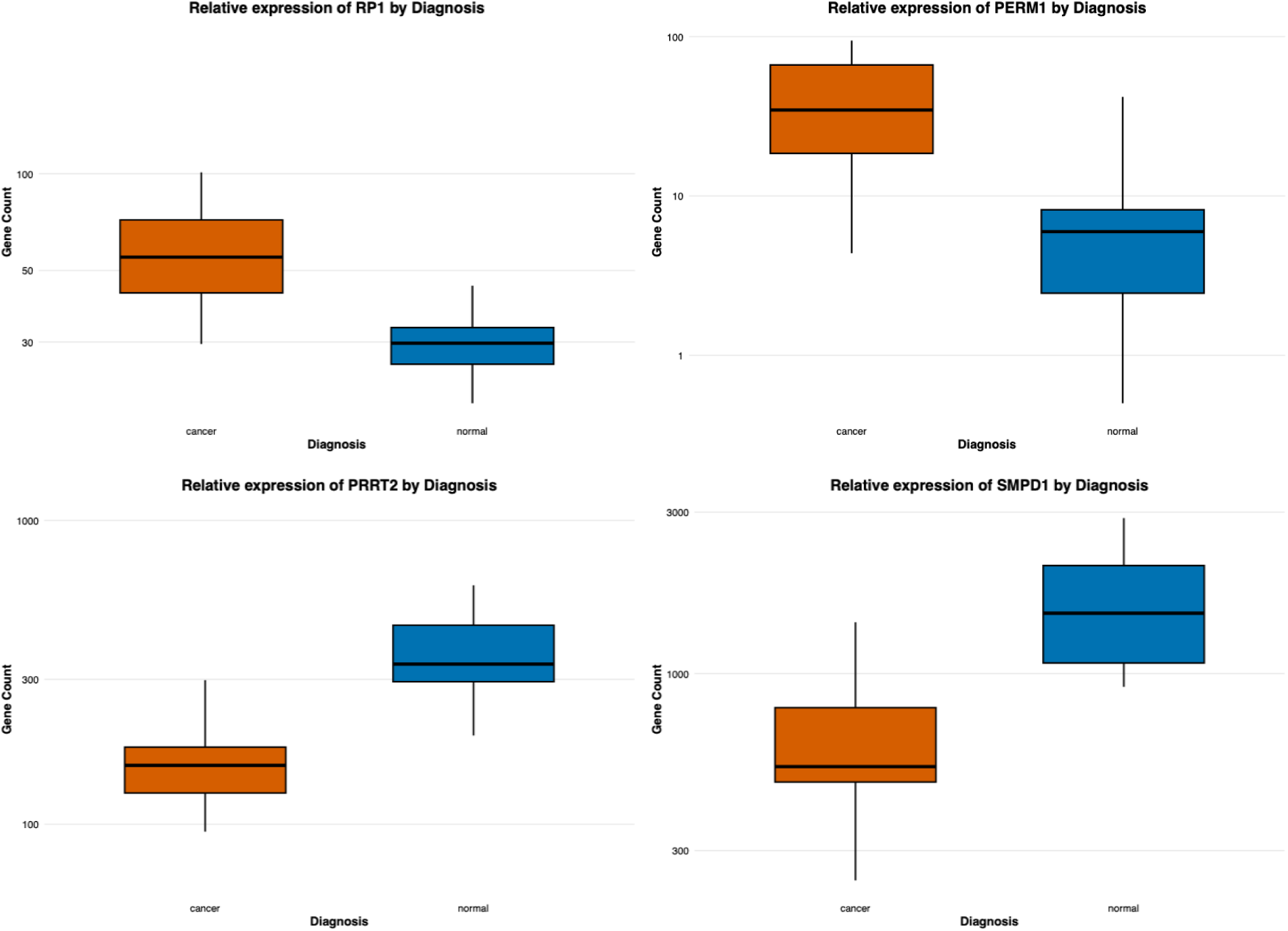
The expression profile in the form of box and whisker plots of the upregulated genes RP1 and PERM1 and the downregulated genes PRRT2 and SMPD1. X-axis represents cancer or normal tissue, Y-axis represents the respective gene counts. The transcription data from the RNA-Seq analysis was used.

### Biomarker identification and testing yields promising novel targets in a four-gene panel

The biomarkers are expected to have DE in cancer vs normal tissues. The significant DE genes as identified from the RNA-seq data (Figure 1B), can be interpreted as potential biomarkers for SA population. In order to assess the accuracy of the genes as potential biomarkers, these genes were subjected to statistical testing to determine their sensitivity, specificity, PPV, and NPV. High PPV and NPV values indicate low false-positive and false-negative rates, respectively, supporting the reliability of these genes as diagnostic biomarkers.

Only protein-coding genes were included, while lncRNAs and miRNAs were excluded, isolating clinically measurable downstream products and the functional effectors of the cancer. Genes with performance values greater than 80% across all metrics were retained. Table 7 presents the results of the manual measurements and filtering, together with gene descriptions obtained from the NCBI Gene database, highlighting strong biomarker performance across multiple candidates, with *SALL4* achieving 100% across all measures.

**Table 7:** Diagnostic parameters of predicted biomarkers (significant DE genes of interest) for clinical utility in CRC.

| Gene | Description | Sens | Spec | PPV | NPV |
|---|---|---|---|---|---|
| *AGAP3* | Involved with GTP binding and possible BRAF fusion [63]. | 99.99% | 92.30% | 99.98% | 80.00% |
| *AMPD2* | Involved in deaminase activity and Notch3 pathway [64, 65]. | 99.99% | 92.30% | 99.98% | 85.70% |
| *ANKRD10* | Expressed in bone marrow and other tissues, linked to canonical Wnt pathway [66]. | 99.99% | 92.30% | 99.98% | 92.30% |
| *SALL4* | Promotes cell proliferation and is a marker for certain types of cancer [67]. | 100% | 100% | 100% | 100% |

Sens = Sensitivity. Spec = Specificity. PPV = Positive Predictive Value, NPV = Negative Predictive Value

Further expression profiling using publicly available datasets along with survival analysis of these genes was performed, with *SALL4* showing upregulation in cancer vs normal samples (Figure 6a) and patients with lower expression of *SALL4* showing extended survival (Figure 6b). Mining of CoReCG database also confirmed higher expression of *SALL4* in tissues derived from cancer patients [58]. Survival anaysis of other genes using KMPlot has been shown in Supplementary information (Figure S12 to S18).

**Figure 6:**
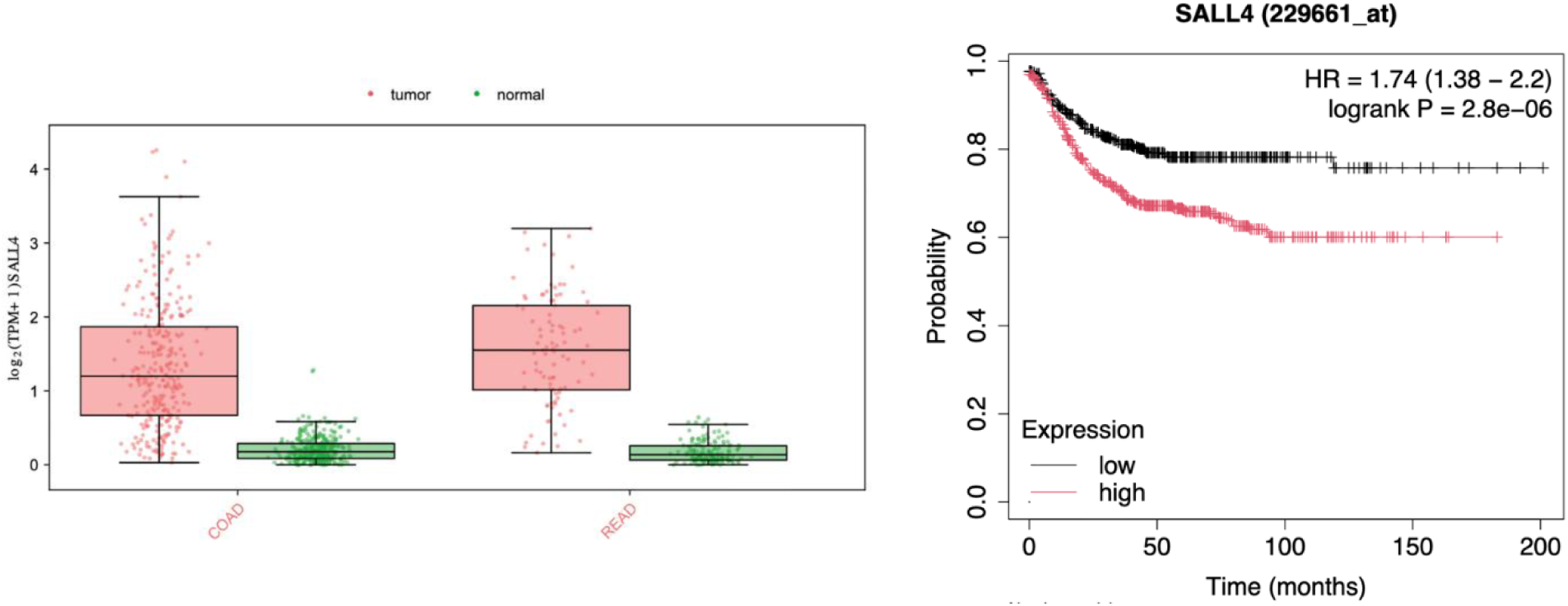
(a) Expression boxplot of SALL4 biomarker candidate gene using TCGA Colon Adenocarcinoma and Rectal Adenocarcinoma (COAD and READ) databases. Boxplot created using GEPIA3 [2S]. Log_2_FC cutoff set at 1 and p<0.05, where each dot represents a samples expression. (b) Kaplan-Meier survival plot using KMPlot [28] showing survival analysis for SALL4 across various studies. The black line indicates survival over time in patients with low expression, while the red line shows the same for high expression.

In addition to the DE genes, the genes harbouring variants of interest (Figure 5) were subjected to manual biomarker testing without performance-based filtering, and the results are presented in Table 8.

**Table 8:** Diagnostic parameters of predicted biomarkers (DE genes harbouring variants of interest) for clinical utility in CRC.

| Gene | Description | Sens | Spec | PPV | NPV |
|---|---|---|---|---|---|
| *SMPD1* | A lysosomal acid sphingomyelinase that converts sphingomyelin to ceramide [68]. | 99.96% | 0 | 99.97% | 0 |
| *PERM1* | Peroxisome receptor coactivator, involved in muscle activity, and mitochondrial biogenesis [69]. | 99.97% | 46.15% | 99.99% | 33.33% |
| *RP1* | Encodes a doublecortin family member, which bind microtubules and regulate microtubule polymerisation [70]. | 99.98% | 46.15% | 99.98% | 50.00% |
| *PRRT2* | Encodes a transmembrane protein, with mice studies suggesting expression in nervous system [71]. | 99.96% | 7.69% | 99.97% | 5.00% |

Sens = Sensitivity. Spec = Specificity. PPV = Positive Predictive Value, NPV = Negative Predictive Value

Further expression information is shown in Supplementary Information (Figures S19 to S21).

The capability of *SALL4* to act as a biomarker is further enhanced with the survival analysis plot as well as the tissue expression plot (Figure 6*a&b*) showcasing distinct correlation with expression and patient outcome. Additionally, *ANKRD10* and *AMPD2* (Table 7) exhibit superior performance (85%+) across all evaluated metrics and represent promising candidates for future diagnostic and prognostic validation (see expression and survival analysis in Supplementary Figures S13 and S14). The genes harbouring variants of interest, *SMPD1*, *PERM1*, *RP1*, and *PRRT2* (Table 8), demonstrate high sensitivity and positive predictive value, indicating strong performance in CRC detection but limited utility for distinguishing non-CRC conditions. These genes hence illustrate the interplay between genomic variation and transcriptomic regulation, underscoring the value of integrative multi-omics approaches.

## Discussion

The primary aim of this study was to generate novel CRC sequencing data from SA patients in order to better characterise the molecular biology of the disease within a historically underrepresented population. Most large-scale CRC sequencing efforts have been conducted in European and North American cohorts, where recurrent alterations in key oncogenic drivers, particularly APC in the Wnt pathway and KRAS in the MAPK pathway, have been firmly established [72–74]. However, growing evidence suggests that the mutational landscape of CRC varies across groups of differing ancestries, with African-American patients displaying higher KRAS mutation frequencies and a broader spectrum of *APC* variants compared to Caucasian and European cohorts [75, 76]. These observations underscore the importance of population-specific genomic studies and provide a strong rationale for the present investigation.

The *APC* variants in Table 5 show multiple inactivating mutations, much like a similar pattern that is observed in intestinal stem cells, where tumorigenesis is driven by *APC* mutations that are beneficial to the tumour survival, with additional research showing multiple *APC* mutations and polyclonality of the tumour assists with overcoming fitness barriers [77, 78]. The pathogenic variants in this study, while some are present in genome databases, their frequency across populations remains unknown, suggesting their rarity. Furthermore, of the truncating variants the c.1620dup and c.4666dup variants have been found in European, Indian and Latin American familial adenomatous polyposis (FAP) populations [79–81]. The c.2547_2550del is a rare germline variant, also associated with FAP in Danish and German populations [82, 83]. The variants, to the authors knowledge, no known literature confirming their presence in African populations. These findings reinforce the central role of Wnt dysregulation in CRC pathogenesis while highlighting population-specific genetic diversity.

Consistent with previous reports, an inverse relationship between *APC* and *BRAF* alterations was observed in MSS tumours [10]. In this cohort, MSS tumours exhibited relatively few *BRAF* mutations (three variants: one synonymous and two missense), in keeping with prior studies demonstrating increased *APC* mutation frequency in tumours with low microsatellite instability. A recent whole-exome sequencing study of sub-Saharan African CRC patients reported an *APC* mutation frequency of 49%, closely mirroring the 55% observed here, yet substantially lower than frequencies reported in TCGA and UK cohorts (70% and 79%, respectively) [8, 13, 84]. Another recent study in Uganda observed similar heterogeneity in *APC* mutations [85]. Conversely, West African cohorts display markedly lower frequencies of Wnt pathway alterations in MSS tumours (37%) [86]. In contrast, *APC* mutation frequencies were higher in patients of African descent than in European patients, but this pattern shifted with increasing age, such that mutation rates were higher among European patients at older ages [10]. When comparing early-onset to average-onset CRC in American patients, it was found that among the top somatically altered genes, *APC* ranked highest with 78.7% mutation rate across both groups [87]. Collectively, these findings point to substantial regional and ancestral heterogeneity in CRC molecular architecture across African populations.

While genomic alterations establish the oncogenic foundation of tumour development, cancer progression is ultimately shaped by the TME, one of the main drivers and initiators of cancer [88]. Established literature shows that stromal remodelling and altered extracellular matrix mechanics promote tumour cell proliferation, invasion, and epithelial–mesenchymal transition, while hypoxia-driven angiogenesis supports tumour expansion and selects for more aggressive phenotypes [89]. In parallel, tumours actively reprogram the immune microenvironment to favour immunosuppression through the recruitment of regulatory and myeloid-derived cells, enabling immune evasion and sustained growth [90]. Transcriptomic profiling in this study revealed a pronounced enrichment of inflammatory and immune-related pathways in SA CRC when compared to normal tissue samples. It is proposed here that the higher inflammatory TME, with the contributors discussed below, participates in the earlier and more aggressive onset of CRC in African patients. Upregulated pathways included cytokine signalling, TNF, IL-17, and NF-κβ signalling, indicating a highly inflamed TME. These findings align with growing recognition that successful tumours rely not only on intrinsic genomic instability and driver mutations, but also on extrinsic interactions with stromal, immune, and vascular components [91].

Emerging evidence has further implicated viral infections as contributors to oncogenesis, a concept supported by the enrichment of the KEGG pathway “viral protein interaction with cytokine and cytokine receptors” in the cancer vs normal cohort (Supplementary Figure S11). This pathway interfaces with multiple cytokine gene families, including CCL, CXCL, interleukins, and TNF. Viral-associated carcinogenesis is well established in cervical cancer (human papillomavirus) and hepatocellular carcinoma (hepatitis viruses) [92, 93], and viral infections in this patient population may similarly contribute to a chronically inflamed microenvironment through immune pathway manipulation. Persistent immune activation can promote oncogenic processes such as oxidative stress, angiogenesis, cellular proliferation, dysregulated signalling, and cytokine-mediated tissue remodelling [94, 95]. GO biological process networks further suggest the presence of a cytokine storm–like phenotype within the TME. In parallel, upregulation of the p53 signalling pathway likely reflects a cellular attempt to counterbalance oncogenic stress induced by chronic inflammation [96]. Public datasets such as TCGA COAD have similarly described immune-enriched tumour phenotypes, supporting the potential relevance of immunotherapy in subsets of CRC patients [97, 98].

Notably when looking at the effect of treatment, chemotherapy-treated tumours exhibited further amplification of inflammatory signalling, whereas untreated tumours showed a more moderate inflammatory profile. This suggests that therapeutic intervention may exacerbate immune activation within the tumour milieu, potentially contributing to treatment response. The inflammatory phenotype observed here is particularly striking and may reflect a defining feature of CRC biology in SA patients. It may also present a therapeutic opportunity, combining chemotherapies with immunotherapies (chemoimmunotherapy) as a treatment regime, which has proved successful in clinical studies for cancers such as non-squamous cell lung carcinoma, and triple negative breast cancer [99, 100]. Despite these notable clinical successes, they have largely not been accompanied by detailed mechanistic investigations or the identification of robust predictive biomarkers [101]. Recent investigation into enhancing immunotherapy effectiveness in MSS tumours have also yielded promising results by combining chemotherapy with immune checkpoint inhibitors, and proposals such as altering the gut microbiome and using probiotic vaccines to reprogram the TME continue to be researched [102]

Beyond immune–tumour interactions, CRC progression also disrupts the complex neuro-immune networks of the gastrointestinal tract, seen across the different contrasts. The gut–brain axis (GBA) represents a bidirectional communication system linking the central nervous system (CNS), enteric nervous system (ENS), microbiota, and immune and endocrine pathways [103]. Uniquely, the ENS retains autonomous regulatory capacity, controlling peristalsis, blood flow, and immune surveillance [104]. In this study, GO biological process mapping revealed downregulation of neuronal and neuromuscular pathways, suggesting progressive atrophy of the ENS within tumour regions [105]. As tumours remodel the ECM, they secrete proteolytic enzymes such as matrix metalloproteinases (MMPs), degrading surrounding tissue and releasing growth factors that facilitate invasion and metastasis [106]. Progressive tumour infiltration leads to destruction of enteric innervation, with degeneration of nerve fibres and atrophy of the submucosal and myenteric plexuses adjacent to tumour tissue [105]. This neurodegeneration may contribute directly to gastrointestinal dysfunction, consistent with clinical histories in which digestive symptoms often precede CRC diagnosis. Furthermore, chemotherapy-induced peripheral neuropathy (CIPN) may exacerbate neuronal injury, providing a plausible explanation for the observed downregulation of membrane action potential regulation, muscle contraction, and locomotion-related pathways in Figure 3 [107]. Together, these findings highlight CRC as a disease that disrupts not only tissue architecture but also fundamental biological function.

At the genomic level, WGS identified 64,504 coding variants, of which 23,026 are catalogued in ClinVar and 14,356 in COSMIC. Prioritisation of “Pathogenic” and “Likely Pathogenic” variants yielded 24 candidates of interest. Notably, only four of these genes exhibited significant DE in the transcriptomic data of cancer vs normal: upregulated *PERM1* (rs13302983 and rs13303355, present in all samples) and *RP1* (rs1437782, present in 50% of samples), and downregulated *PRRT2* (rs11150573, present in all samples) and *SMPD1* (c.138_143del, present in 50% of samples). This limited concordance between genomic variation and transcriptomic dysregulation likely reflects the complexity of regulatory mechanisms, inter-individual genetic variability, chemotherapy effects, and the influence of non-coding regions on gene expression.

Furthermore, WGS analysis provided insight to eight novel, functional variants shown in Table 6. Notably, the *CARD11* and *TRIMc4B* variants are linked to inflammatory response, supporting the hypothesis that SA CRC carcinogenesis is linked to inflammation. The genes *PITRM1* and *SLCcA1c* are associated with the nervous system, providing evidence for the impact between the gut and brain axis.

Building on these molecular insights, biomarker analysis evaluated eight candidate genes (*AGAP3*, *AMPD2*, *ANKRD10*, *SALL4*, *SMPD1*, *PERM1*, *RP1*, and *PRRT2*) using local patient data and published literature. Only *SALL4* has previously been implicated in CRC [108], with the remaining genes representing novel candidate markers in South African CRC.

Finally, CMS classification [109] revealed that 30% of tumours mapped to CMS4 and 30% to CMS3, indicating a predominantly mesenchymal and metabolically driven tumour phenotype. This contrasts with TCGA cohorts, which show greater representation of CMS1 and CMS2 subtypes [110, 111]. CMS4 tumours are characterised by epithelial– mesenchymal transition, stromal infiltration, angiogenesis, and poor prognosis, while CMS3 tumours display metabolic dysregulation. The predominance of these subtypes in this cohort aligns closely with the observed inflammatory microenvironment, ECM remodelling, neurodegeneration, and metabolic reprogramming. Together, these features may contribute to the poorer clinical outcomes reported in African CRC patients and suggest biologically distinct disease mechanisms in SA populations. This study further enriches the molecular level understanding of CRC in SA patients and adds to the knowledge of potential models of tumourigenesis (Table 9) prevalent among different populations.

**Table 9:** Comparison of potential models of tumourigenesis between European and African patients.

| **European CRC model** | **African CRC model** |
|---|---|
| Epithelial, mutation-driven disease [112, 113] | Systems-level, microenvironment-driven disease [114] |
| Dominated by APC/KRAS/TP53 [113] | Shaped by inflammation, infection, stromal biology [114] |
| CMS1–CMS2 enriched [110, 115] | CMS3–CMS4 enriched [116] |
| Predictable driver architecture | High allelic heterogeneity |

In summary, this study provides one of the first integrated genomic and transcriptomic characterisations of SA CRC. The findings reveal a disease driven not only by canonical oncogenic pathways but also by profound immune activation, microenvironmental remodelling, neuro-immune disruption, and population-specific genetic variation. These results highlight the importance of population-representative cancer genomics and support the development of precision oncology strategies tailored to African patients.

### Limitations and Shortcomings

This study has several limitations that should be considered when interpreting the findings. All patient samples were obtained from Wits Donald Gordon Medical Centre, a private hospital in Johannesburg. As a result, the cohort may not fully represent the broader SA population, particularly individuals treated in the public healthcare sector. Socioeconomic and healthcare access disparities may therefore introduce selection bias and limit the generalisability of the findings.

The study is further constrained by the limited number of patient samples, which restricts statistical power and limits the ability to perform robust subgroup analyses across clinical, pathological, and molecular features. In addition, incomplete historical clinical records and limited longitudinal follow-up data precluded detailed correlation of molecular profiles with treatment response, disease progression, and survival outcomes.

From a technical perspective, computational resource constraints restricted the scale of multi-omics integration and limited the exploration of non-coding regulatory variation, structural variants, and epigenetic mechanisms that may further contribute to CRC pathogenesis in this population. Due to the limited computation capacity at the time of study, individual VCF files were stored for future research.

Despite these limitations, this study establishes a foundational genomic and transcriptomic resource for SA CRC. Importantly, it has catalysed ongoing efforts to expand patient recruitment through public hospitals in Johannesburg to increase cohort diversity and improve population representation. Future studies will incorporate larger patient cohorts, comprehensive clinical annotation, and longitudinal follow-up to enable outcome-driven molecular stratification.

In addition, planned wet-lab validation studies using patient-derived tumour organoids will provide functional characterisation of candidate driver mutations and biomarkers. The organoid establishment protocol has already been published [117], and these models will enable further biological explorations, drug response profiling and mechanistic validation, strengthening the translational relevance of the findings.

## Conclusion

This study represents a seminal contribution towards addressing the long-standing sequencing research gap in SA CRC. We establish, for the first time, a comprehensive pipeline for patient recruitment, biospecimen processing, high-throughput sequencing, and integrative bioinformatic analysis tailored specifically to SA CRC.

Through robust RNA sequencing, we identified key DE genes across chemotherapy-treated and untreated tumours, with *AGAP3*, *AMPD2*, *ANKRD10*, and *SALL4* demonstrating exceptional diagnostic performance (average sensitivity of 99.9%, specificity of 94.22%, positive predictive value of 99.9%, and negative predictive value of 89.5%). These findings nominate a new class of highly accurate transcriptomic biomarkers for CRC detection in South African patients. Furthermore, functional analyses revealed a highly inflammatory TME driving carcinogenesis, with links to viral influences, and notable disruption to neuronal signalling linking to the GBA.

Integration of WGS further generated a first-of-its-kind catalogue of genomic variation in this cohort, revealing pathogenic and likely pathogenic variants with direct transcriptomic impact. In particular, *SMPD1*, *PERM1*, *PRRT2*, and *RP1* emerge as promising multi-omics biomarkers, bridging genomic variation with functional dysregulation. Furthermore, eight novel variants in genes *KDMcA*, *C2CD2*, *TAF2*, *SLCcA1c*, *OR5M10*, *PITRM1*, *TRIMc4B*, *CARD11* are reported with links to inflammation and neural processes.

Collectively, these findings substantially expand the genomic and transcriptomic resources available for African cancer research and provide critical insight into the molecular architecture of CRC in an underrepresented population. More broadly, this work underscores the importance of population-specific cancer genomics and lays the foundation for precision oncology strategies tailored to African patients. Continued expansion of these efforts will be essential to translate these discoveries into improved diagnostic, prognostic, and therapeutic interventions for CRC in SA and beyond.

## Data Availability

The data supporting the conclusions of this article is available upon request.

## Declarations

### Ethics approval and consent to participate

Ethics approval was granted by the Human Research Ethics Committee Medical of the University of the Witwatersrand (M201233). Informed written consent from patients was granted for the use of their collected tissue and publication of research findings. The experimental protocols conform to the Declaration of Helsinki.

### Availability of data and materials

The data supporting the conclusions of this article is available upon request.

### Research Funding

Financial Research support (No: 129356) from the National Research Foundation (NRF), South Africa to MK and bursary to ZK (MND200503519532) are acknowledged.

### Authors’ contributions

ZK and NA contributed to collection and processing of samples and writing of the manuscript. ZK analysed and interpreted the data and created figures. BB provided the patient samples. CK assisted with genetic sequencing. AC and PM assisted with WGS analysis and interpretation. MK conceptualised the study, interpretation of data, generated funds, supervised, established collaborations for this work. All authors assisted with editing of the manuscript.

### Supplementary Material

Supplementary Information.docx

Supplementary Table.xlsx

