## Supplementary file for "Transcriptomic and Genomic Sequencing of South African Colorectal Cancer Patients Reveal Population-Specific Insights and Variants"

### SUPPLEMENTARY INFORMATION

#### Quality Control: GC content and Mean Quality Scores for the RNA-seq samples

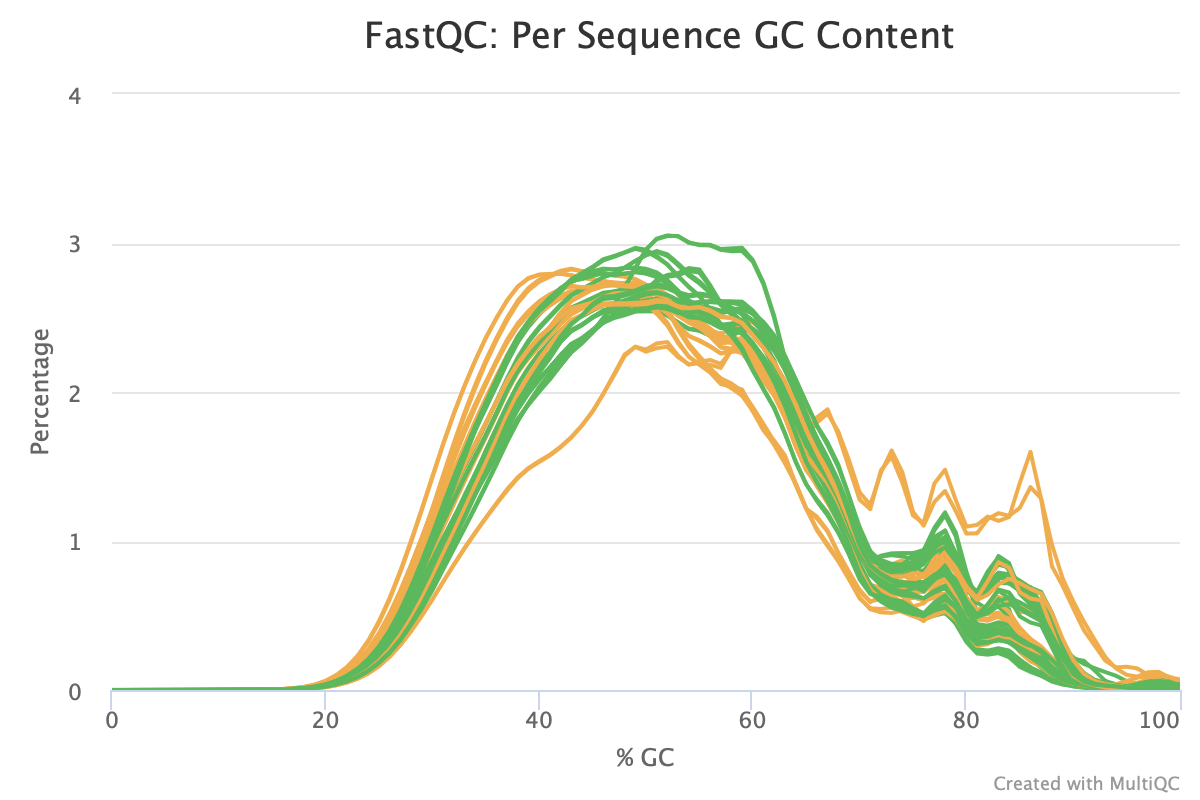

Figure S1: The per sequence GC content of the RNA-seq samples.

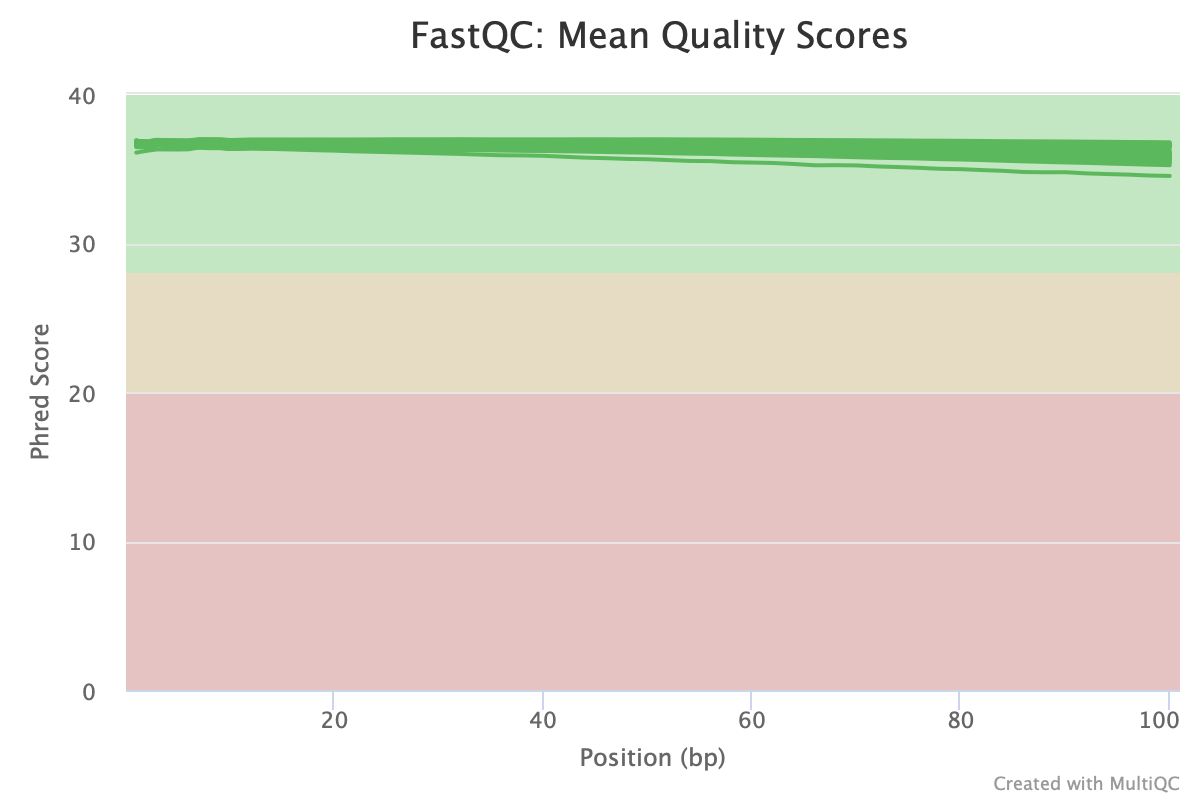

Figure S2: The mean quality scores for each RNA-seq sample

#### Differential Expression Results for Chemotherapy Treated and Untreated Samples

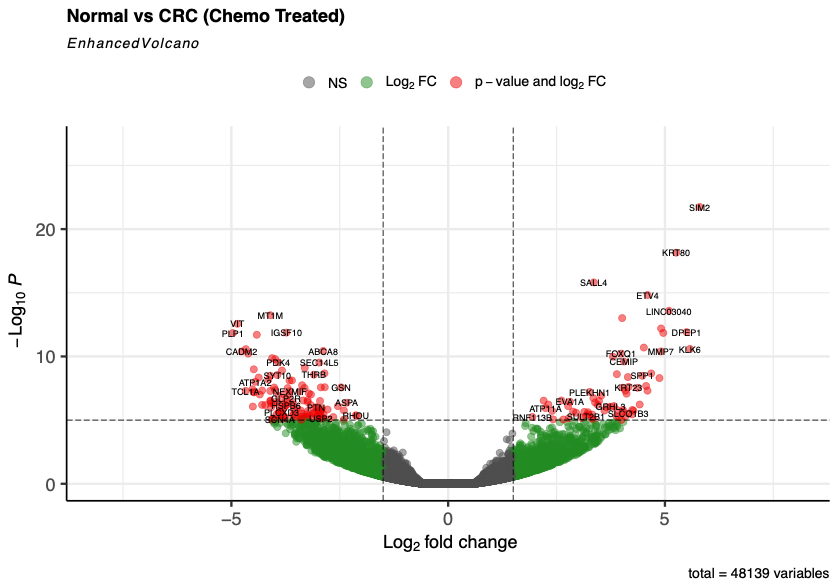

Figure S3: Volcano Plot for Chemotherapy Treated Samples showing differentially expressed genes. The x-axis indicates the log_2_ FC scale from negative to positive, and the y-axis represents the log_10_ P value indicating significance. The graph is split into sections with dotted lines using the thresholds set. Each gene is represented by a dot, green dots representing genes with no significant expression, green dots representing genes that meet the log FC threshold of significance, blue dots representing genes that meet the p-value threshold for significance and red dots representing genes that meet both thresholds

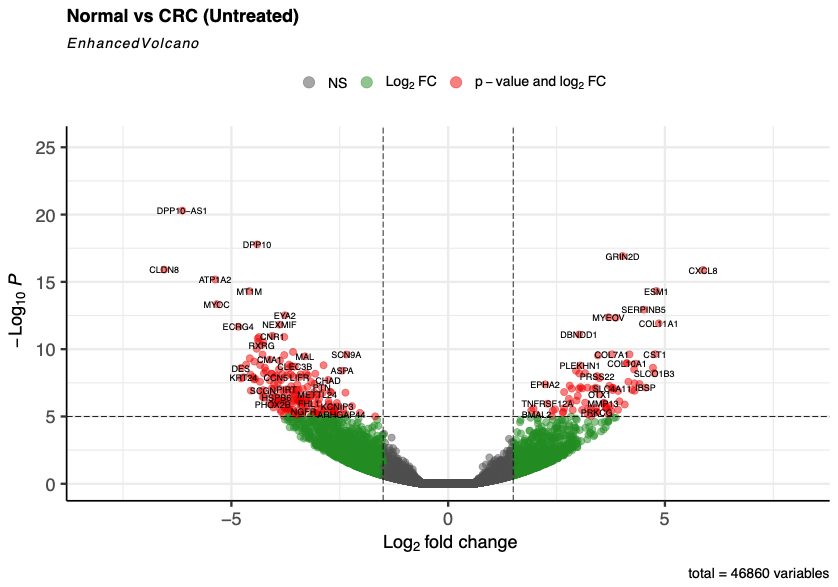

Figure S4: Volcano Plot for Chemotherapy Untreated Samples showing differentially expressed genes. The x-axis indicates the log_2_ FC scale from negative to positive, and the y-axis represents the log_10_ P value indicating significance. The graph is split into sections with dotted lines using the thresholds set. Each gene is represented by a dot, green dots representing genes with no significant expression, green dots representing genes that meet the log FC threshold of significance, blue dots representing genes that meet the p-value threshold for significance and red dots representing genes that meet both thresholds

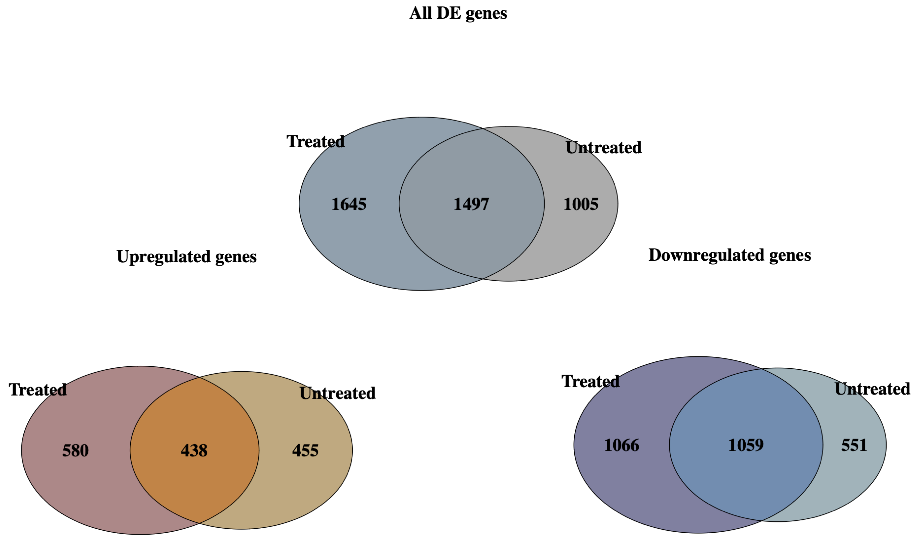

#### Functional Analysis Results For Chemotherapy Treated and Untreated Samples

Figure S5: Venn diagram describing the gene overlap between chemotherapy treated and untreated samples for their respective DE genes (up and down regulated genes).

.The genes were matched to their respective GO Terms, and where genes mapped to multiple a connecting line was formed. Each GO term is represented as a green circle, with brighter circles indicating higher statistical significance and larger circles representing a greater number of mapped genes. The connecting lines between GO terms vary in thickness, ranging from faint grey to thick grey, depending on the degree of gene overlap between terms. This network structure facilitates the identification of functionally related biological processes and their relative significance within the dataset.

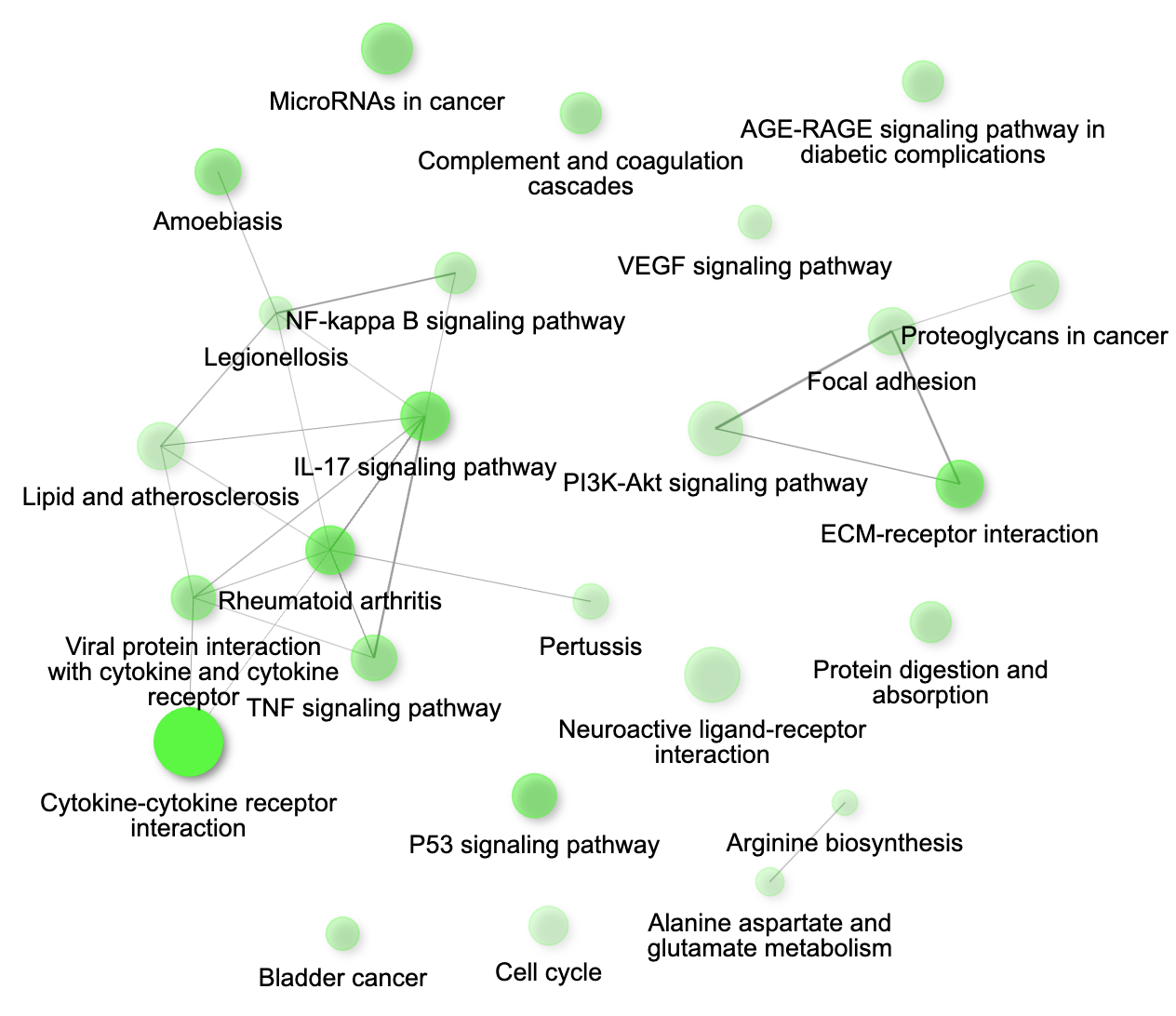

Figure S6: Upregulated KEGG pathways in the chemotherapy untreated samples presented as a network

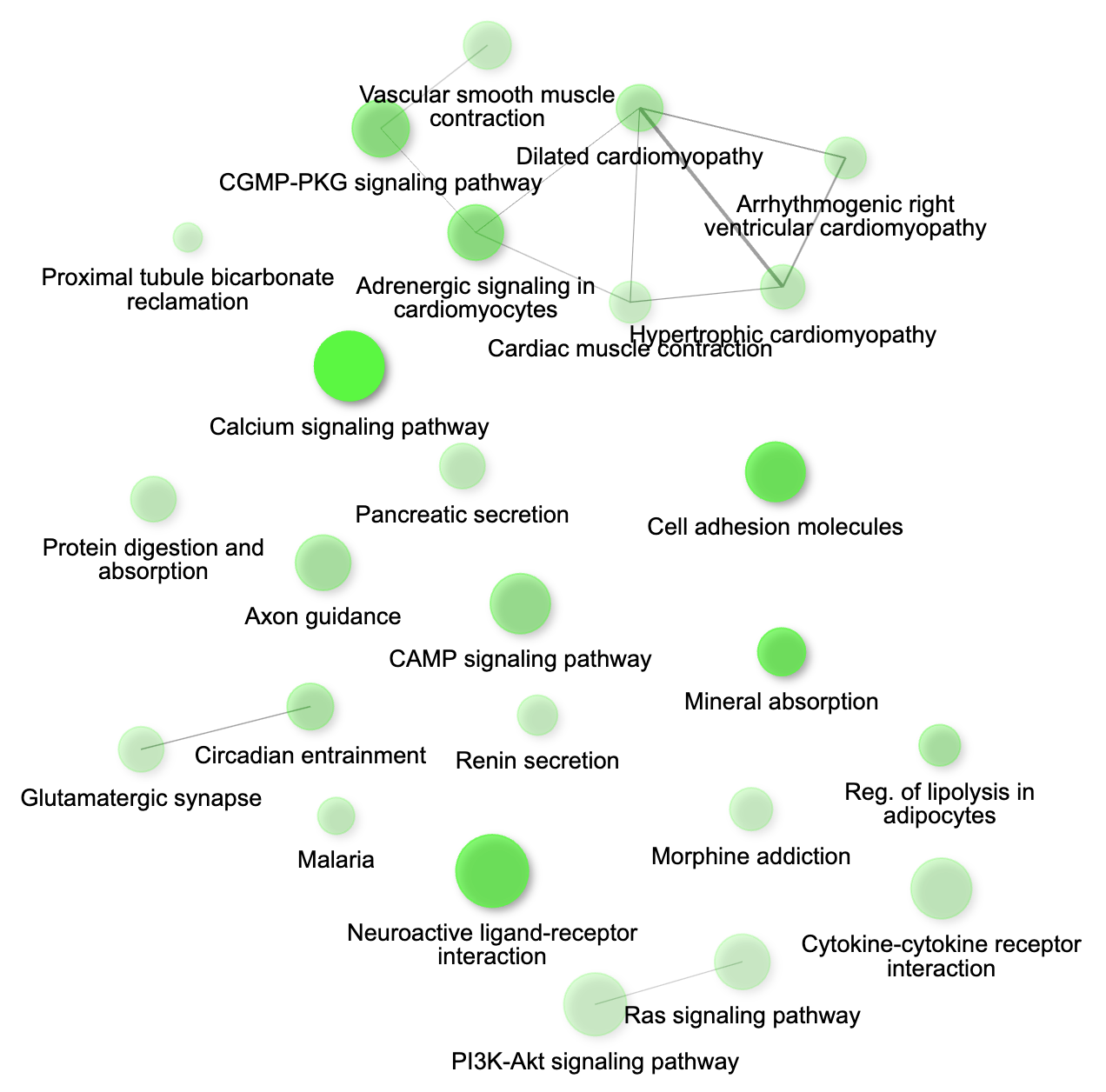

Figure S7: Downregulated KEGG pathways in the chemotherapy untreated samples

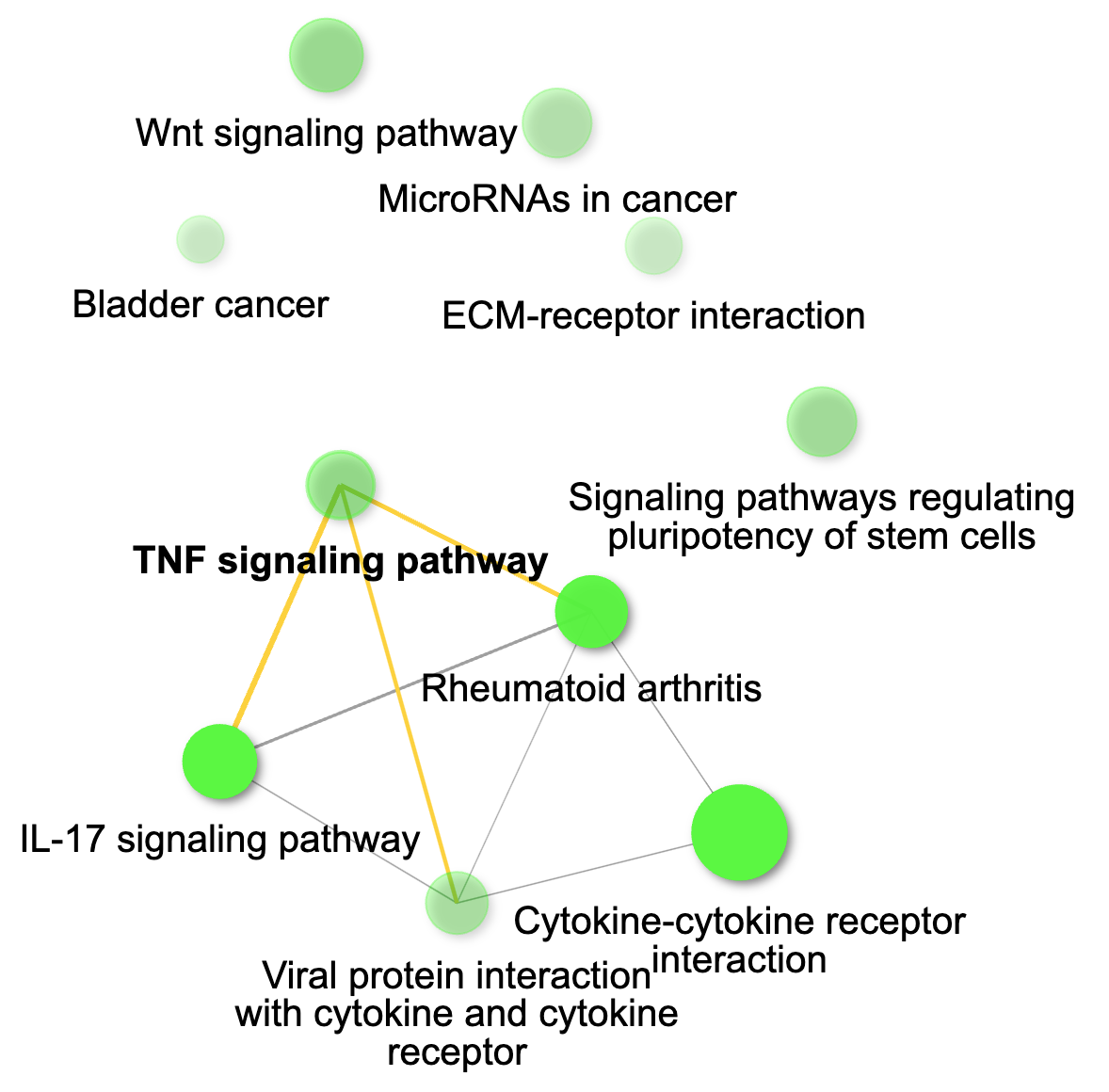

Figure S8: Upregulated KEGG pathways in the chemotherapy treated samples

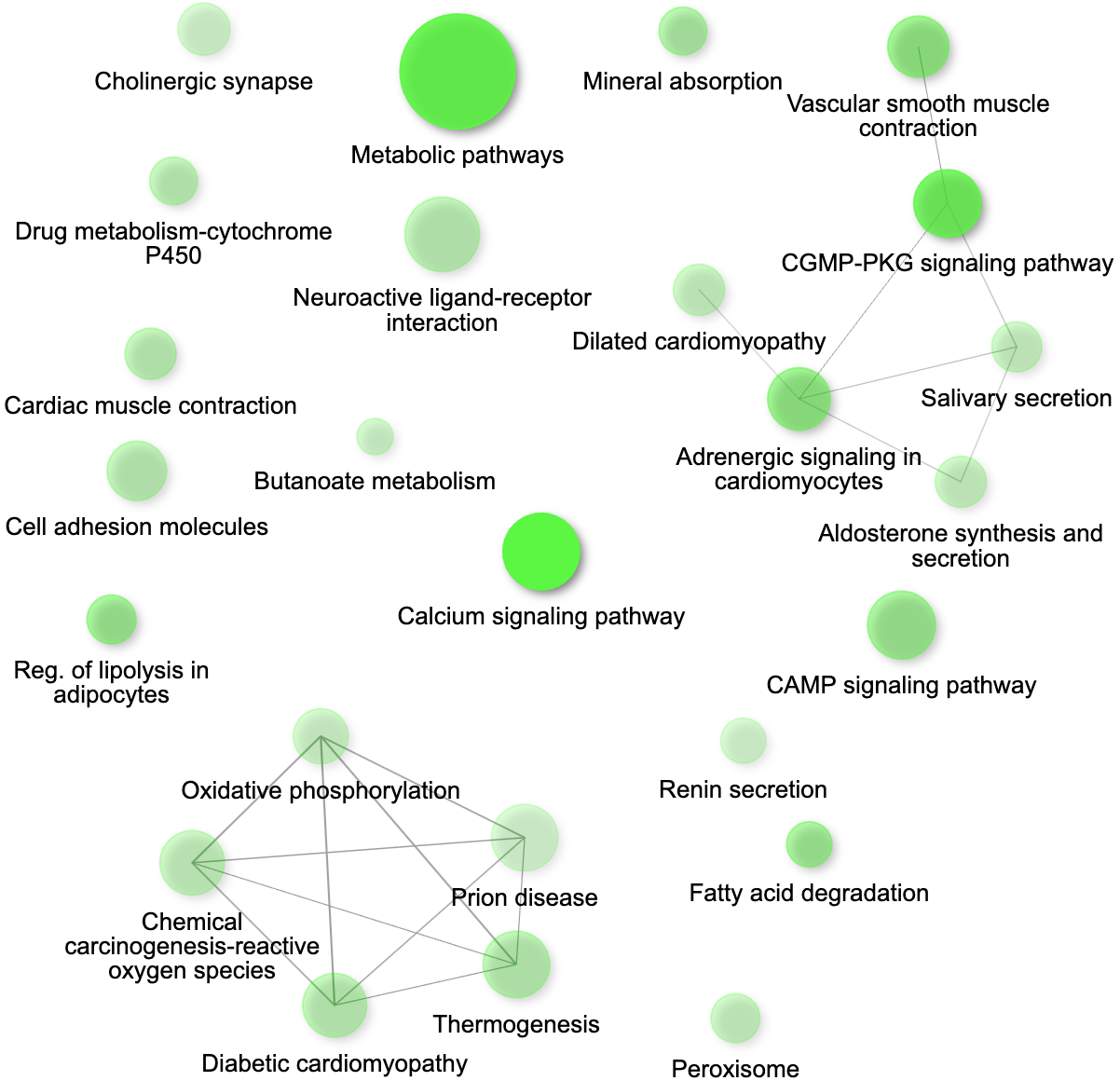

Figure S9: Downregulated KEGG pathways in the chemotherapy treated samples

**A**

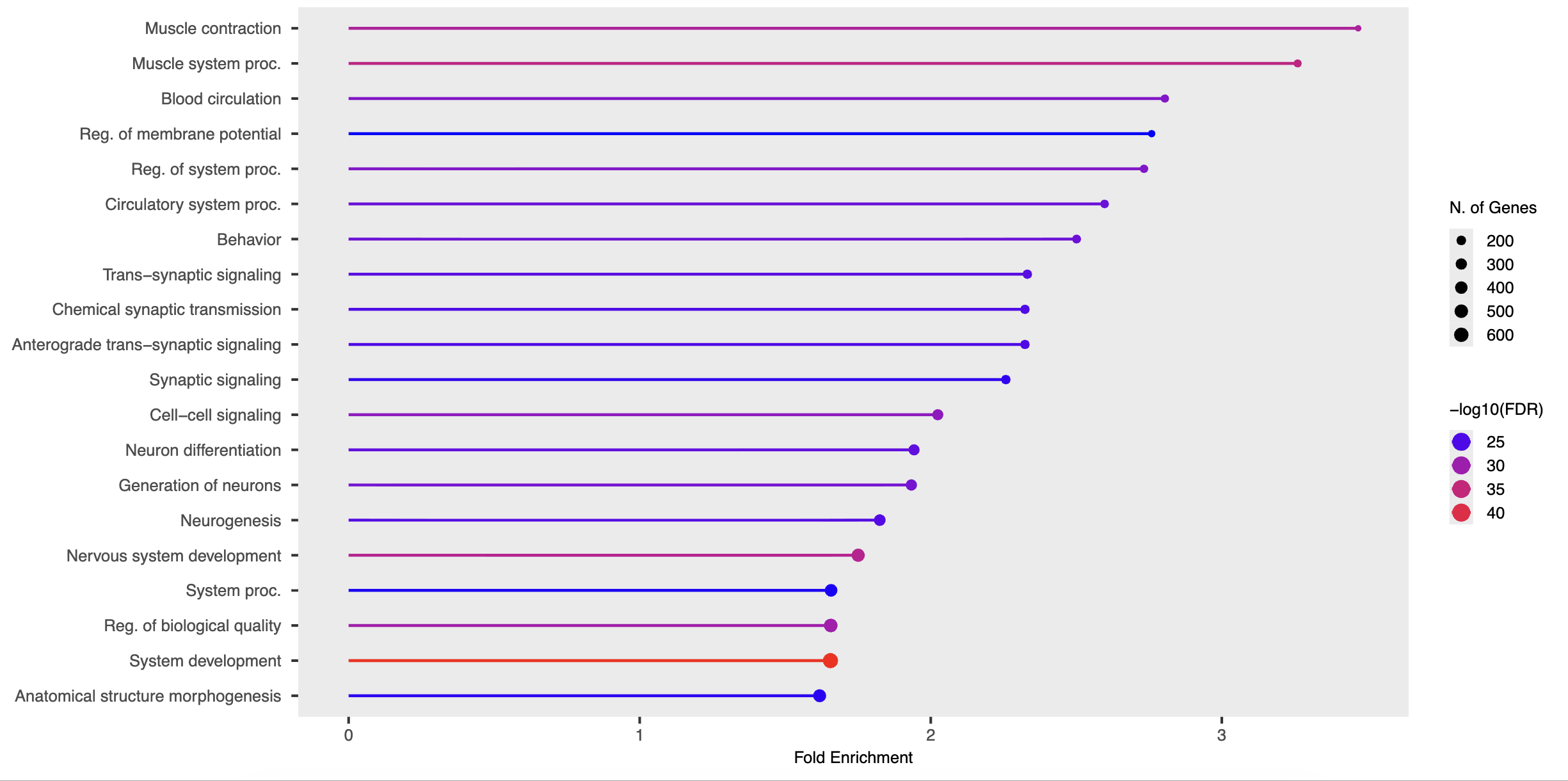

**B**

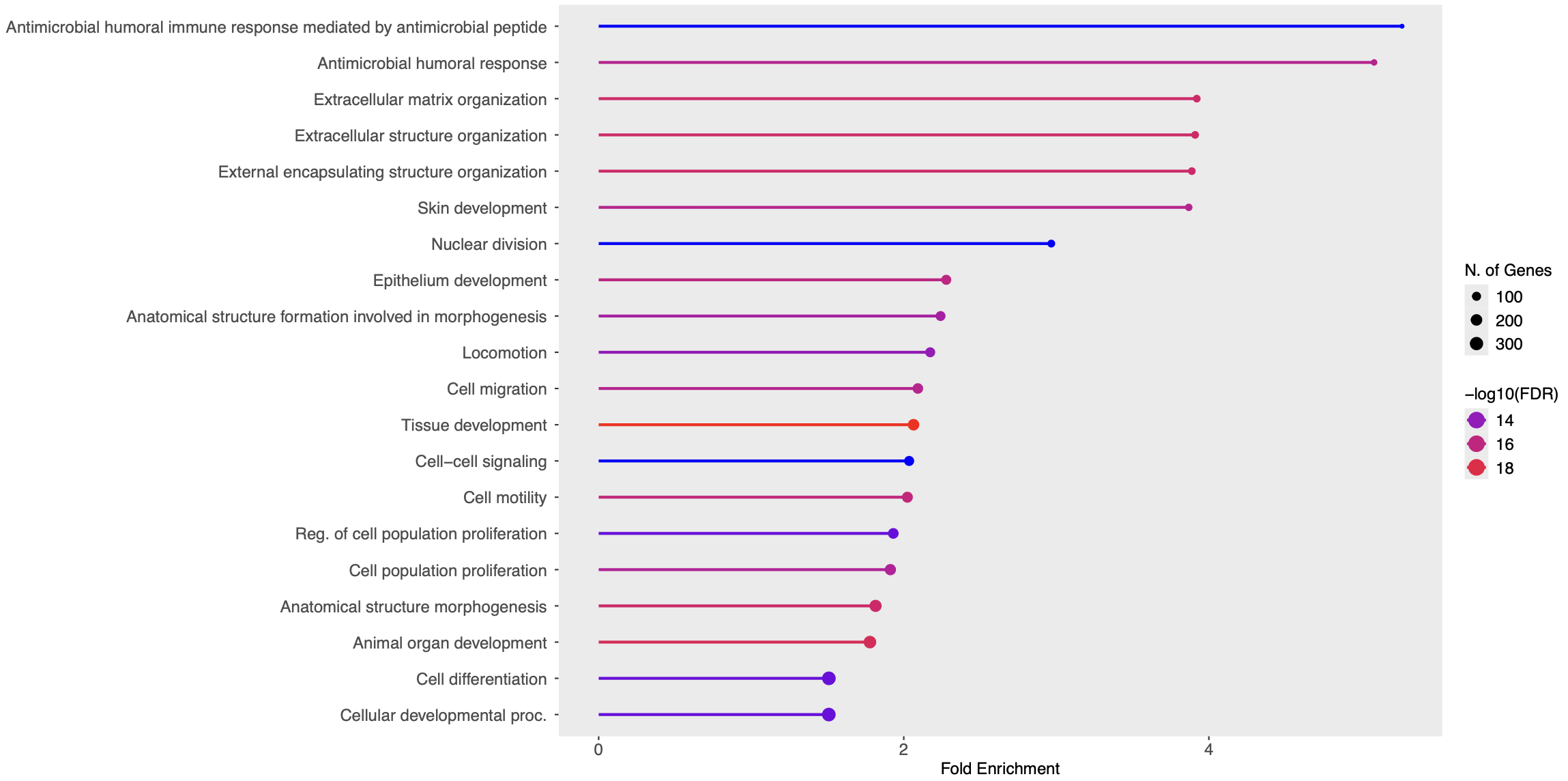

Figure S10: (A) The top 20 enriched GO Processes for the up and (B) downregulated genes. The colour of each bar indicated the false discovery rate, the length of the bar indicates the fold enrichment (x-axis), while the size of the black dot indicated the number of genes involved in the biological process.

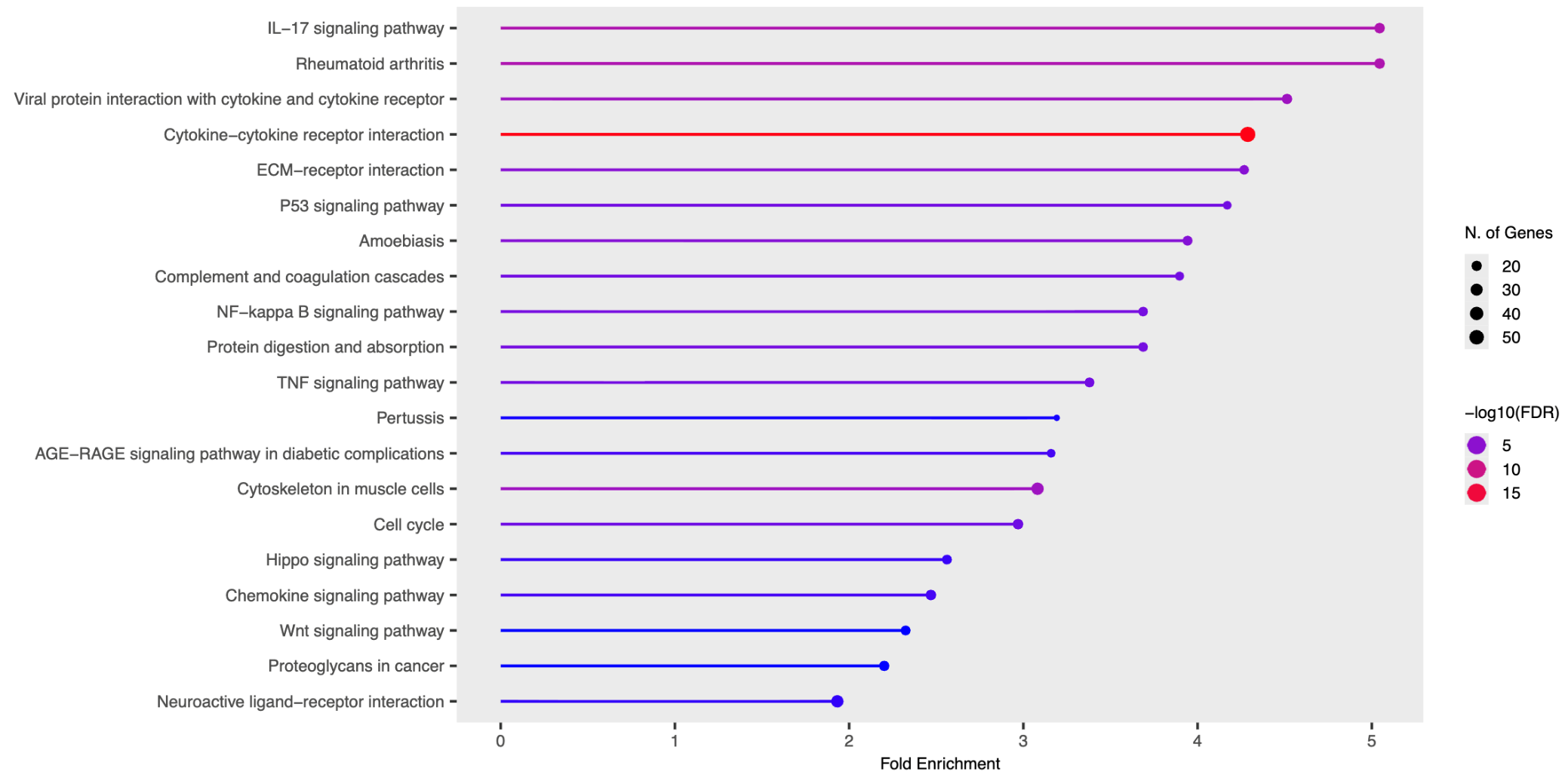

**A**

*Inflammatory TME*

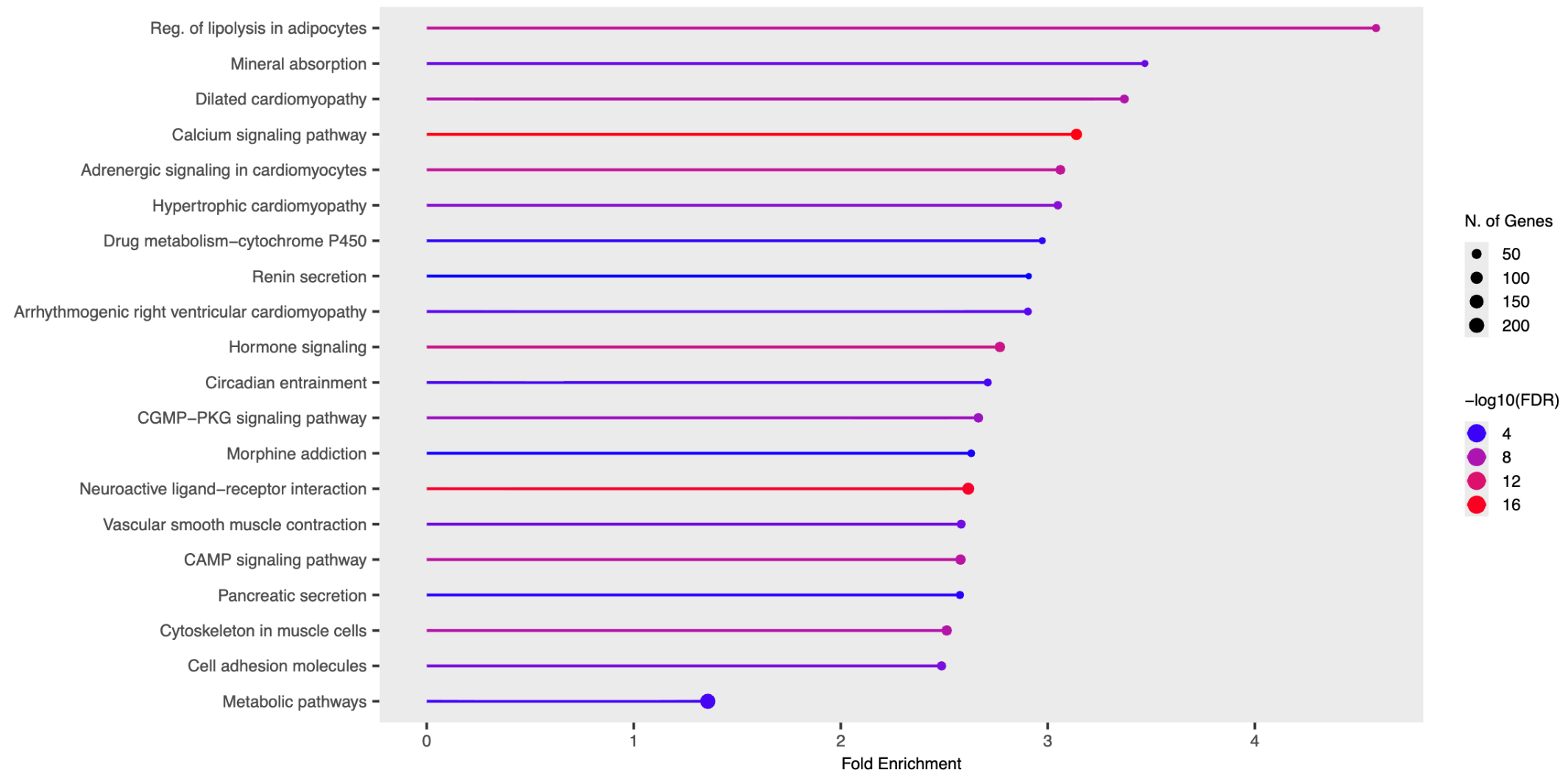

Figure S11: The top 20 enriched KEGG pathways for the (A) up and (B) downregulated genes. The colour of each bar indicated the false discovery rate, the length of the bar indicates the fold enrichment (x-axis), while the size of the black dot indicated the number of genes involved in each KEGG pathway.

**B**

#### Additional WGS Results

Table S1: Variant types present in the main coding regions

| Variant Type | Count |
| --- | --- |
| FRAME_SHIFT_DEL | 82 |
| FRAME_SHIFT_INS | 71 |
| IN_FRAME_DEL | 187 |
| IN_FRAME_INS | 175 |
| MISSENSE | 11335 |
| NONSENSE | 84 |
| NONSTOP | 17 |
| SILENT | 13068 |
| START_CODON_DEL | 2 |
| START_CODON_INS | 3 |
| START_CODON_SNP | 12 |

#### Biomarker Analysis – Survival Analysis using KMPLOT

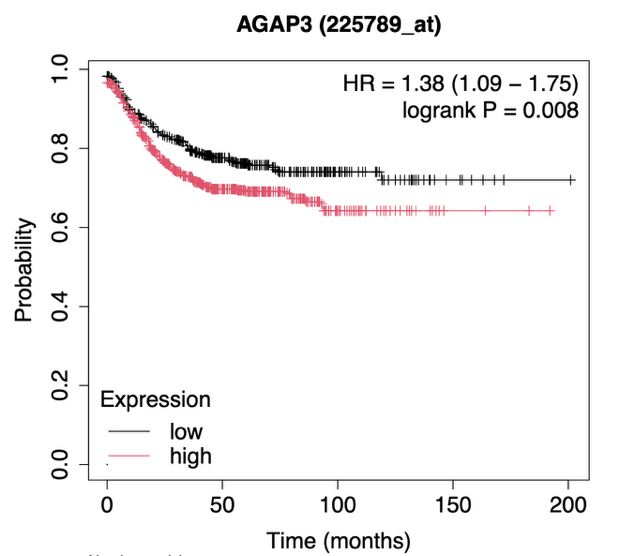

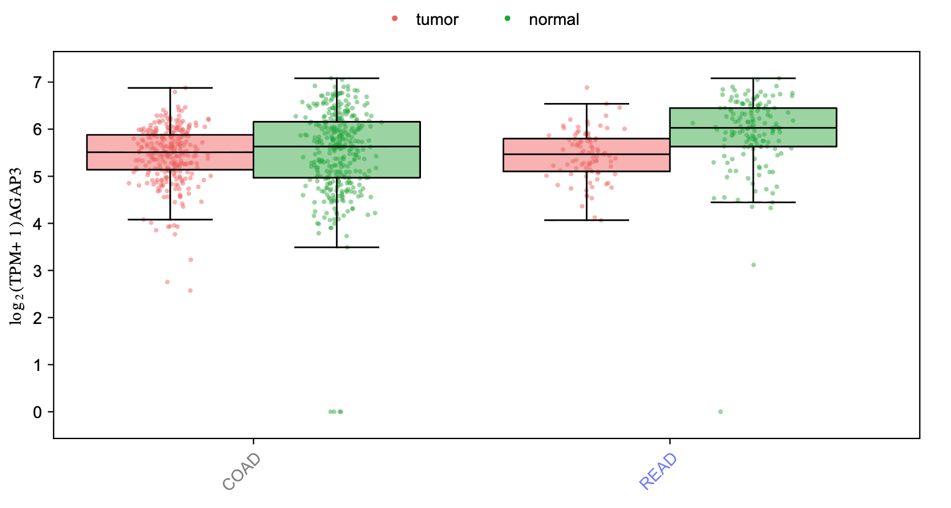
Kaplan-Meier plots created using KMPlot using Affymetrix gene data for colon cancer studies.

Figure S12: (a) Expression boxplot of AGAP3 biomarker candidate gene using TCGA Colon Adenocarcinoma and Rectal Adenocarcinoma (COAD and READ) databases. Boxplot created using GEPIA3. (b) Kaplan-Meier survival plot using KMPlot showing survival analysis for AGAP3 across various studies.

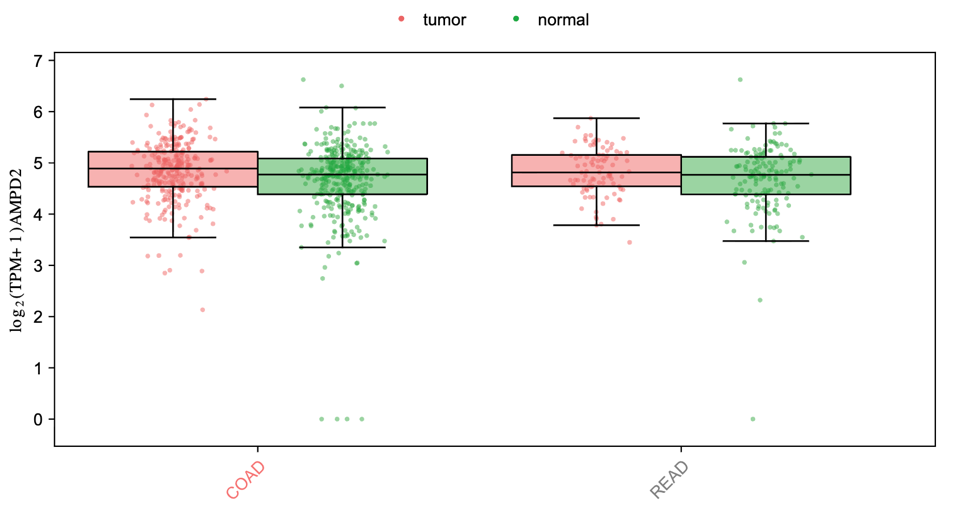

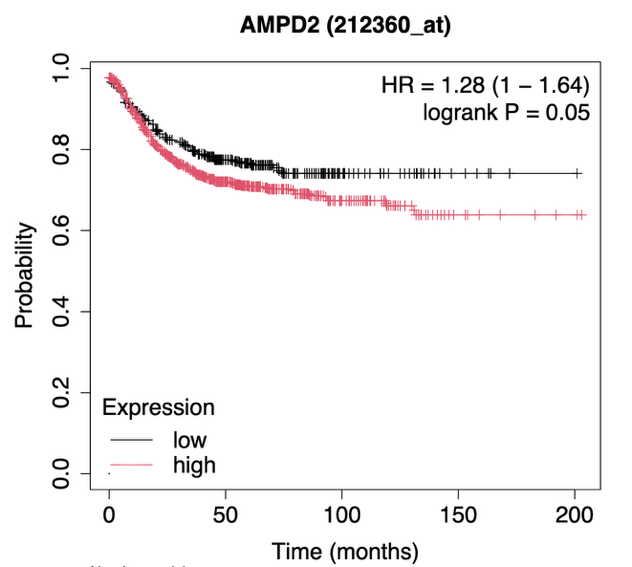
*Figure S13: (a) Expression boxplot of AMPD2 biomarker candidate gene using TCGA Colon Adenocarcinoma and Rectal Adenocarcinoma (COAD and READ) databases. Boxplot created using GEPIA3. (b) Kaplan-Meier survival plot using KMPlot showing survival analysis for AMPD2 across various studies.*

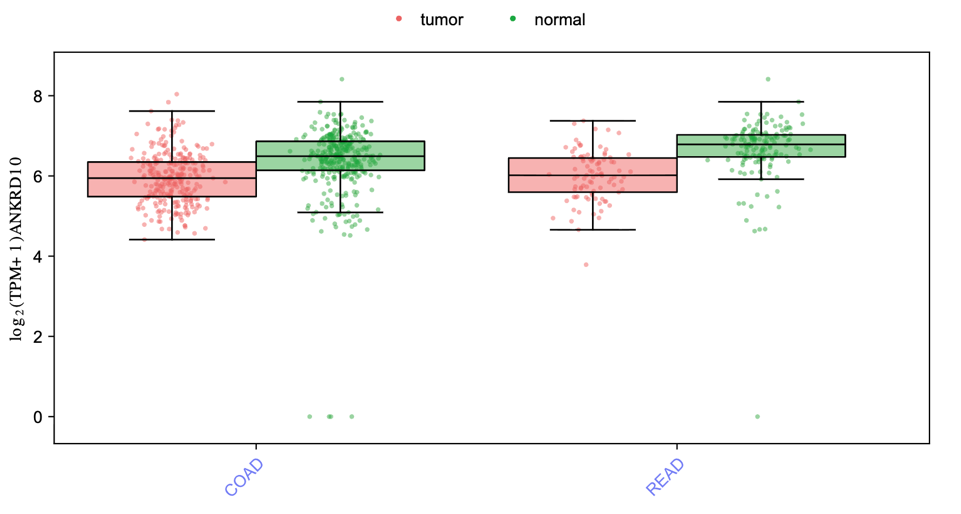

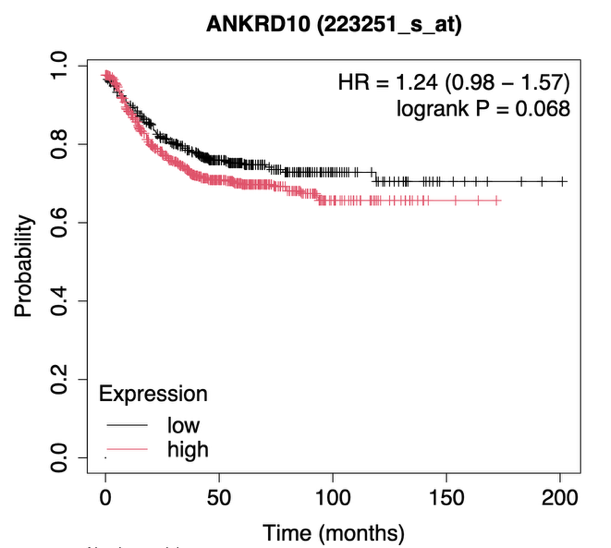

Figure S14: (a) Expression boxplot of ANKRD10 biomarker candidate gene using TCGA Colon Adenocarcinoma and Rectal Adenocarcinoma (COAD and READ) databases. Boxplot created using GEPIA3. (b) Kaplan-Meier survival plot using KMPlot showing survival analysis for ANKRD10 across various studies.

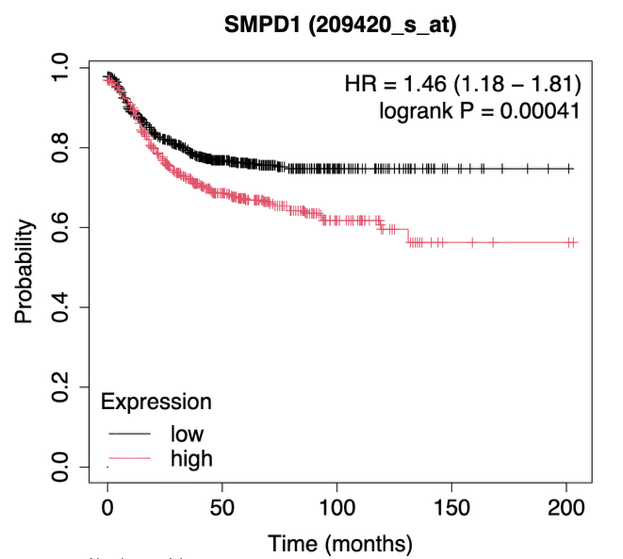

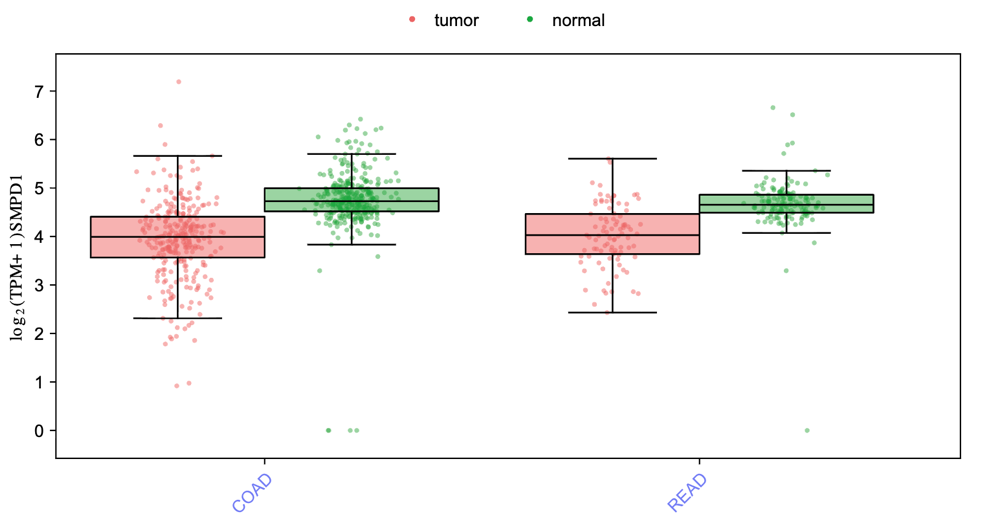

Figure S15: (a) Expression boxplot of SMPD1 biomarker candidate gene using TCGA Colon Adenocarcinoma and Rectal Adenocarcinoma (COAD and READ) databases. Boxplot created using GEPIA3. (b) Kaplan-Meier survival plot using KMPlot showing survival analysis for SMPD1 across various studies.

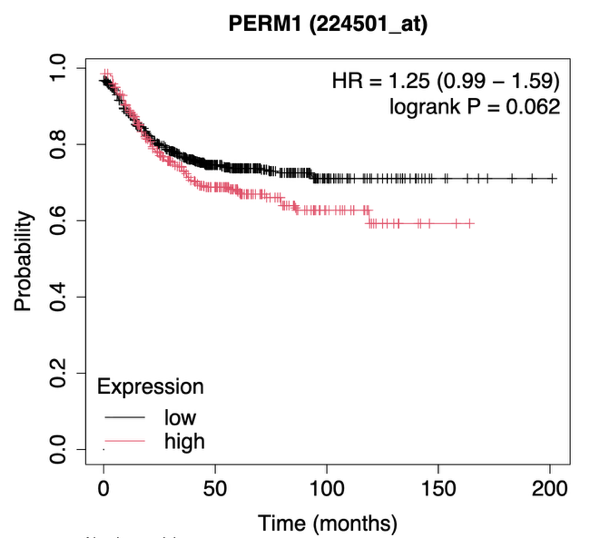

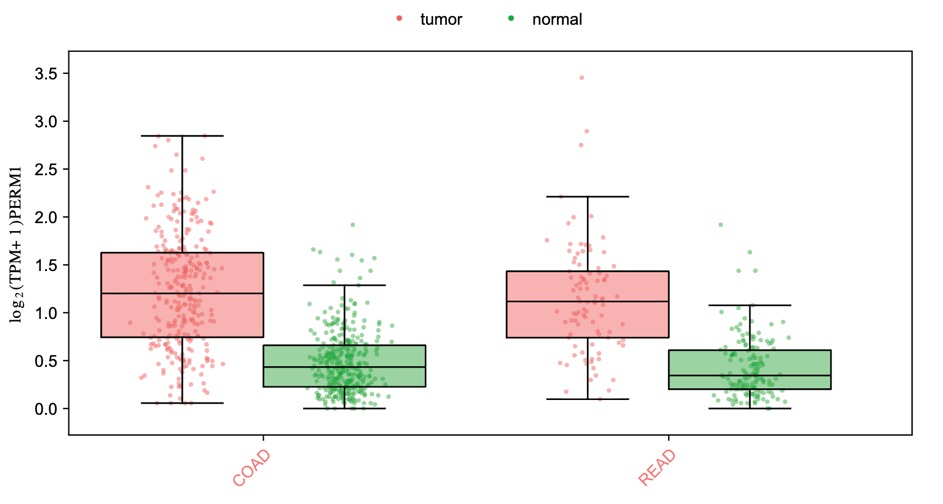

Figure S16: (a) Expression boxplot of PERM1 biomarker candidate gene using TCGA Colon Adenocarcinoma and Rectal Adenocarcinoma (COAD and READ) databases. Boxplot created using GEPIA3. (b) Kaplan-Meier survival plot using KMPlot showing survival analysis for PERM1 across various studies.

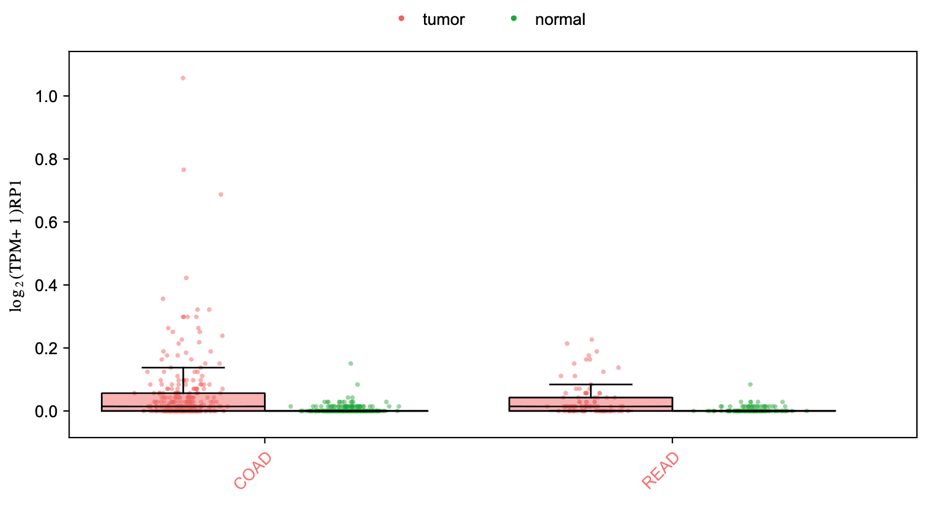

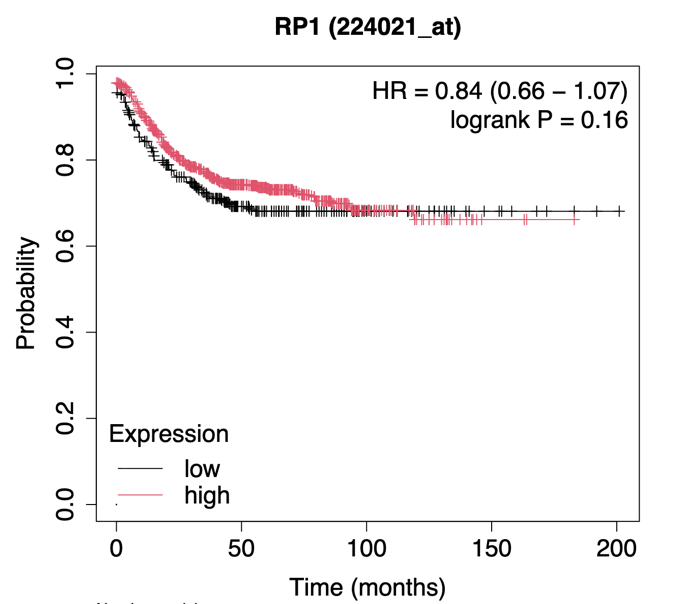

Figure S17: (a) Expression boxplot of RP1 biomarker candidate gene using TCGA Colon Adenocarcinoma and Rectal Adenocarcinoma (COAD and READ) databases. Boxplot created using GEPIA3. (b) Kaplan-Meier survival plot using KMPlot showing survival analysis for RP1 across various studies.

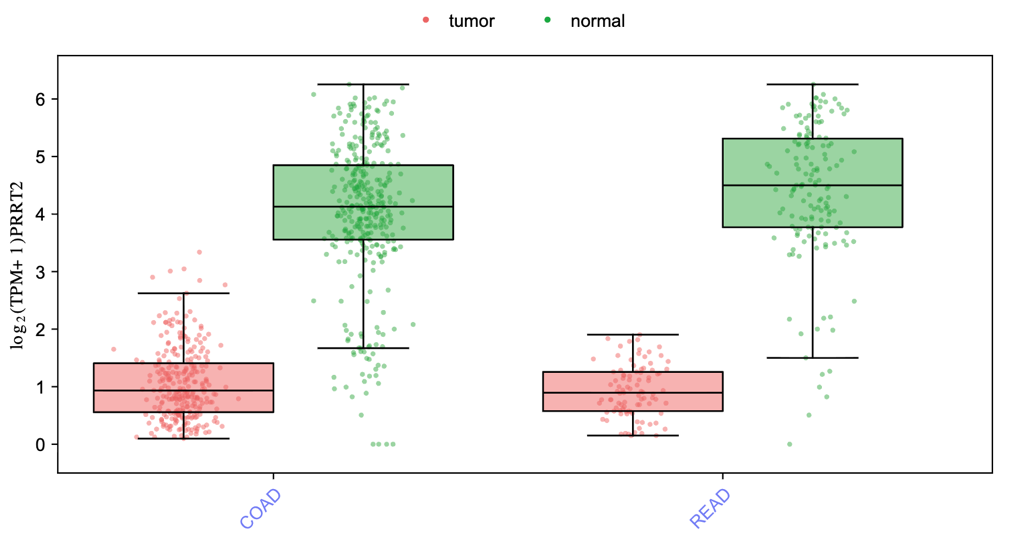

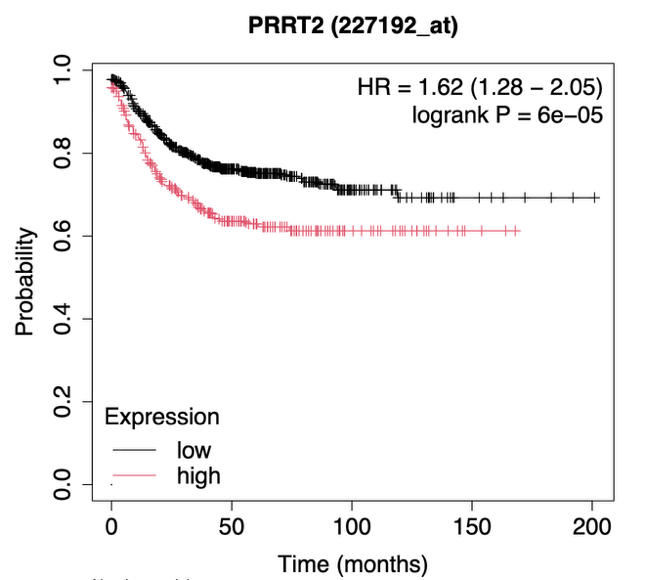

Figure S18: (a) Expression boxplot of PRRT2 biomarker candidate gene using TCGA Colon Adenocarcinoma and Rectal Adenocarcinoma (COAD and READ) databases. Boxplot created using GEPIA3. (b) Kaplan-Meier survival plot using KMPlot showing survival analysis for PRRT2 across various studies.

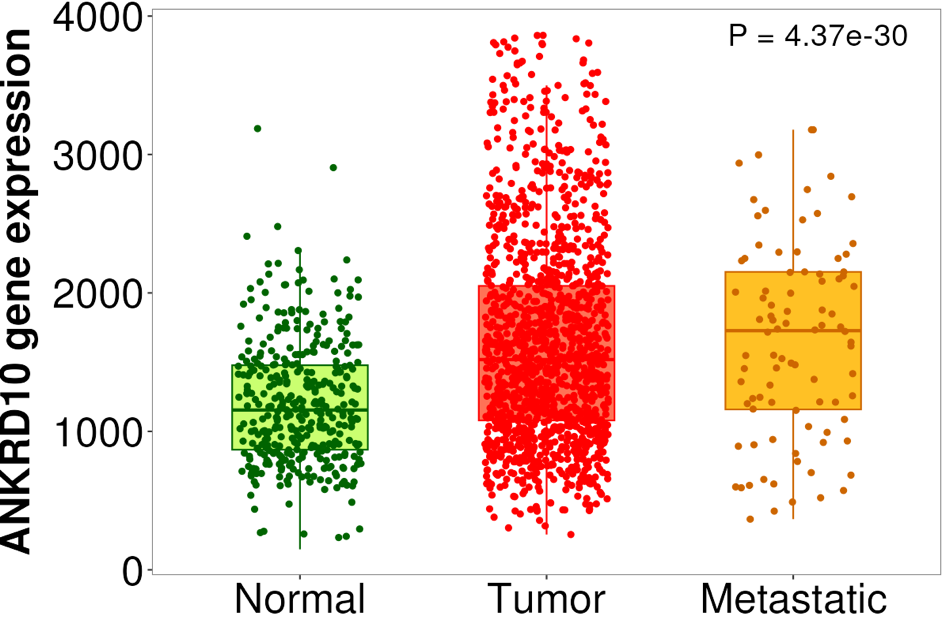

Figure S19: Box and whisker expression plot across normal, tumour and metastatic cells for ANKDRD10. Created using TNMPlot and TCGA COAD and READ Data.

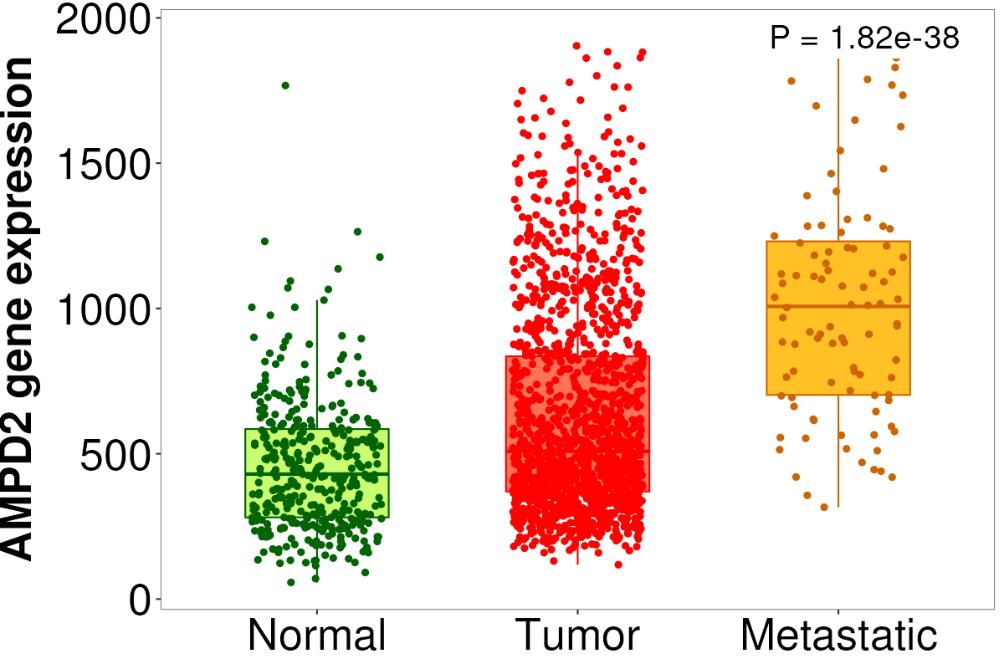

Figure S20: Box and whisker expression plot across normal, tumour and metastatic cells for AMPD2. Created using TNMPlot and TCGA COAD and READ Data.

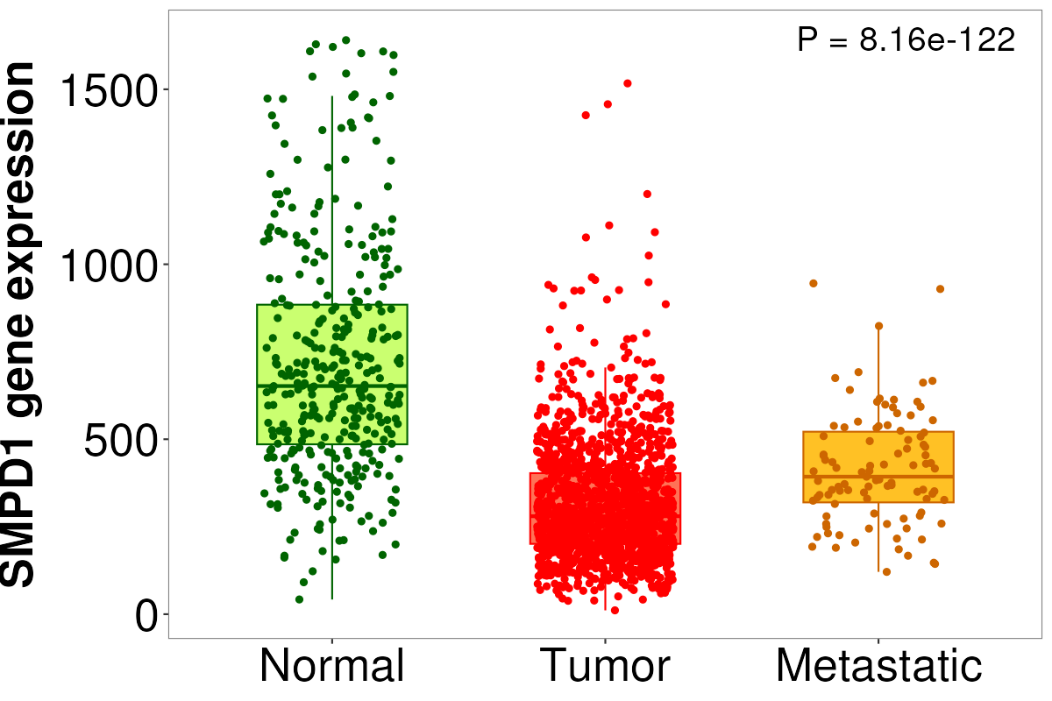

Figure S21: Box and whisker expression plot across normal, tumour and metastatic cells for SMPD1. Created using TNMPlot and TCGA COAD and READ Data.
